# Metabolic traits and the risk of head and neck cancer: a systematic review and meta-analysis

**DOI:** 10.1101/2023.09.19.23295793

**Authors:** Alexander J Gormley, Charlotte Richards, Francesca Spiga, Alex Whitmarsh, Emily Gray, Jo Hooper, Barry G Main, Emma E Vincent, Rebecca C Richmond, Julian PT Higgins, Mark Gormley

**Affiliations:** Bristol Dental School, Faculty of Health Sciences, University of Bristol; Bristol Dental Hospital, University Hospitals Bristol and Weston NHS Foundation Trust; School of Dentistry, Cardiff University; Population Health Sciences, Bristol Medical School, University of Bristol; MRC Integrative Epidemiology Unit, Population Health Sciences, Bristol Medical School, University of Bristol; Library and information services, University Hospitals Bristol and Weston NHS Foundation Trust; School of Cellular and Molecular Medicine, Bristol Medical School, University of Bristol

**Author notes:** Joint first authors (Alexander J Gormley, Charlotte Richards).

## Abstract

**Introduction:** The overall incidence of head and neck cancer (HNC) continues to rise, despite a decline in smoking rates, particularly within developed countries. Obesity and related metabolic traits have been attributed to a growth in cancer rate, so further exploration of these risk factors is warranted in HNC. A comprehensive systematic review and meta-analysis was conducted in order to obtain the most precise observational estimates between metabolic trait exposures and risk of HNC.

**Methods:** A search strategy was developed with an information and content specialists. Multiple databases including Cochrane Library, OVID SP versions of Medline, EMBASE, pre-prints and the grey literature were searched. The primary outcome for included studies was incident HNC and exposures included obesity defined by body mass index (BMI), type 2 diabetes, dyslipidaemia, and hypertension, using pre-specified definitions. A combined risk effect across studies was calculated using both fixed and random-effects meta-analysis. Heterogeneity was assessed between studies using the Cochrans Q and I^2^ statistical tests. The ROBINS-E preliminary tool was used to assess the bias in each included result.

**Results:** The search generated 7,316 abstracts, of these 197 full text articles were assessed for eligibility and 36 were included for full qualitative and quantitative synthesis. In the analysis of 5 studies investigating the association between obesity and incidence of HNC, there was an overall RR of 1.06 (95%CI (0.76, 1.49), *P* heterogeneity <0.024, I^2^= 73.2%) using a random-effects model. 6 studies reported on the association between type 2 diabetes and incidence of HNC, with an overall RR of 1.13 (95%CI (0.95, 1.34), *P* heterogeneity= <0.0001, I^2^= 80.0%) using a random-effects model. An increased risk of hypertension was consistent across HNC subsites, with the strongest association found in the larynx (RR= 1.17, 95%CI (1.10, 1.25), *P* heterogeneity= 0.186, I^2^= 37.7%). For dyslipidaemia, only 2 studies were available for meta-analysis in the laryngeal subsite, with some evidence of an increased risk association of both low high-density lipoprotein (RR= 1.12, 95%CI (1.07, 1.18), *P* heterogeneity= 0.103, I^2^= 62.5%) and high triglyceride levels (RR= 1.10, 95%CI (1.05, 1.15), *P* heterogeneity= 0.319, I^2^= 0.0%)) using random-effects models. Over 80% of studies were judged to be at Very High or High risk of bias using the ROBINS-E tool.

**Conclusion:** Despite individual studies suggesting an association between BMI and HNC, limited effect was demonstrated in this meta-analysis. There was evidence of an association between hypertension and dyslipidaemia on incident HNC, however caution is required due to the high levels of heterogeneity recorded in these studies. Observational associations are susceptible to confounding, bias and reverse causality so these results must be interpreted with caution. Future work should include meta-analysing studies separately by geographic region, as this appears to be a clear source of heterogeneity.

## Introduction

Despite the significant reduction in smoking ^1^, particularly in developed countries, the overall incidence of head and neck cancer (HNC) continues to rise ^2, 3^. Given this potentially changing aetiology ^4^, further exploration of other risk factors is warranted to enhance prevention strategies. According to the World Health Organization (WHO), at least 40% of all cancer cases may be preventable, with smoking, alcohol consumption and obesity identified as three of the most important modifiable lifestyle factors ^5^. The growth in cancer attributable to metabolic risk is increasing, with a global attributable fraction due to high body mass index (BMI) at around 3.6%, equating to almost half a million new cases each year ^6 7^. Public health strategies have been unsuccessful in addressing the current obesity epidemic at the population level, which could also result in more overall cancer cases in the years to come ^8^. It has been projected that obesity could even supersede smoking as the primary driver of cancer in the coming decades ^5^.

People living with obesity and associated metabolic traits have an increased risk of developing cancer ^5^, but the epidemiological evidence surrounding HNC is not conclusive ^9,10^. In the largest pooled analysis, obesity defined by higher BMI was associated with a protective effect for HNC in current smokers (hazard ratio (HR)= 0.76, 95%CI (0.71, 0.82), *p* <0.0001, per 5 kg/m^2^) and conversely, a higher risk in never smokers (HR= 1.15, 95%CI (1.06, 1.24) per 5 kg/m^2^, *p* <0.001) ^11^. More recent cohort studies have failed to show a clear association between BMI and HNC ^12–16^. Similarly, a meta-analysis of observational studies investigating type 2 diabetes with oral and oropharyngeal subsites found an increased risk ratio (RR) of 1.15, 95%CI (1.02, 1.29) ^17^. This finding has been supported by more recent independent cohorts ^12,18–20^. Lastly, hypertension (raised systolic blood pressure (SBP) or diastolic blood pressure (DBP)), has been correlated with head and neck cancer risk across multiple studies ^19–23^. There are a number of proposed mechanisms linking these metabolic traits and cancer, with none specifically linked to HNC. In short, hyperglycaemia may provide a substrate for tumour growth or lead to direct DNA mutation. Insulin itself is a potent growth factor, thought to promote proliferation and carcinogenesis both directly and indirectly through insulin like growth factor-1 (IGF-1) ^24–26^. Insulin resistance can lead to a chronic proinflammatory environment and the formation of macrophages which produce mutagenic agents and reactive oxygen species leading to DNA damage ^27^. Any underlying mechanism between hypertension and cancer risk remains unknown ^23^. Lipids are an essential component of the cell wall and so increased cellular uptake of cholesterol has been seen in many cancers, while other malignancies have demonstrated lipid metabolism reprogramming which can help promote a cancer cell phenotype including proliferation, differentiation and metastasis ^28^.

We aimed to conduct the largest, most comprehensive systematic review and meta-analysis to date, in order to obtain the most precise observational association between metabolic traits and risk of HNC. If available, results were stratified by subsite (i.e., oral cavity, oropharynx, hypopharynx, and larynx), with pre-defined subgroup analyses performed to enhance interpretation, and additional sensitivity analysis to improve certainty in the results.

## Methods

### Research Question

‘Do metabolic traits affect the risk of developing head and neck cancer?’

### Eligibility criteria

**Population-** participants over 18 years old, of either sex, from any country or ethnic background.

**Exposures**-1) obesity, 2) type 2 diabetes, 3) hypertension and 4) dyslipidaemia as defined in the study protocol below. These will be collectively described as metabolic traits.

**Comparison-** participants who have not been diagnosed with a metabolic trait(s).

**Outcome-** incident head and neck squamous cell carcinoma (HNC).

### Study Selection

We focused on prospective observational studies reporting the risk of metabolic traits on incident HNC. This was to ensure a direct temporal relationship between the exposure and onset of head and neck cancer. Eligibility criteria are detailed in **Table 1**.

**Table 1.** Study selection criteria.

| Inclusion criteria | Exclusion criteria |
| --- | --- |
| All studies published between January 1966 and November 2021, in the English language.<br>Human participants >18 years old, of either sex or any ethnic background.<br>Observational (e.g., cohort studies) reporting the risk of incident head and neck squamous cell carcinoma.<br>Studies must have original data and report an adjusted odds ratios, risk ratio, hazard ratio (HR), or standardized incidence ratios (SIR) with their 95% CIs (or data to calculate them).<br>Exposures: obesity, type 2 diabetes, hypertension, and dyslipidaemia, with definitions as described in the study protocol.<br>Outcome: head and neck cancer (HNC) diagnosis in a major subsite i.e., oral cavity, oropharynx, larynx or hypopharynx, which may be human papilloma virus (HPV) positive or negative. | The study is not based on incident head and neck squamous cell carcinoma, or contains only prevalent data, meaning a temporal relationship cannot be inferred e.g., case-control.<br>Studies focused on cancer survival or progression.<br>Studies with no original data or effect statistic reported. Continuous results to be discounted if they cannot be interpreted/ converted into categorical e.g., BMI >30.<br>Human studies only, no *in vivo* animal or *in vitro* cell line studies.<br>Studies of head and neck epithelial dysplasia, potentially malignant disease, or carcinoma *in situ*.<br>Squamous cell carcinoma of other sites e.g., nasopharynx, salivary gland, oesophagus, skin, or lung due to differing aetiology, histological subtypes, and risk factors e.g., Epstein Barr Virus. |

### Data sources and search strategy

A systematic search strategy (**Supplementary Table 1**) was formulated by clinicians, scientific researchers and a specialist librarian for Head & Neck Surgery and Medicine. Medical Subject Headings (MeSH) and keywords were iteratively combined with the Boolean operators AND, OR and NOT. Multiple databases were searched from January 1966 – November 2021, including OVID SP versions of Medline and EMBASE, using the formulated search strategy (**Supplementary Table 1**). Pre-print servers *medRxiv* (since June 2019) and *BioRxiv* (since November 2013) were also searched. In addition, the following electronic bibliographic databases were comprehensively searched: EThOS, Google Scholar, Open Grey and ClinicalTrials.gov, to identify articles from the grey literature and conference proceedings using a modified search strategy (**Supplementary Table 2**). References extracted from the full-length articles were reviewed to identify other publications of interest. Duplicate articles were removed using Covidence (*Covidence systematic review software*, Veritas Health Innovation, Melbourne, Australia) ^29^.

To ensure transparent and reproducible methodology, this study has been reported in line with the Preferred Reporting Items for Systematic Reviews and Meta-Analyses (PRISMA) statement and was pre-registered on International Prospective Register of Systematic Reviews (PROSPERO) in 2021 (CRD42021250520). The protocol was also accepted for publication in *BMJ Open* ^30^.

### Data screening and extraction

All titles and abstracts were initially screened by two authors (A.G. and C.R.) using Covidence ^29^. Any conflicts were resolved by a third independent reviewer (M.G). A PRISMA flowchart ^31^ representing the study selection process is shown in **Figure 1**. Full text of articles included after title and abstract screening were retrieved and screened by (A.G., C.R and M.G.). Data were then extracted from articles which met the inclusion criteria (**Table 1**).

**Figure 1.**
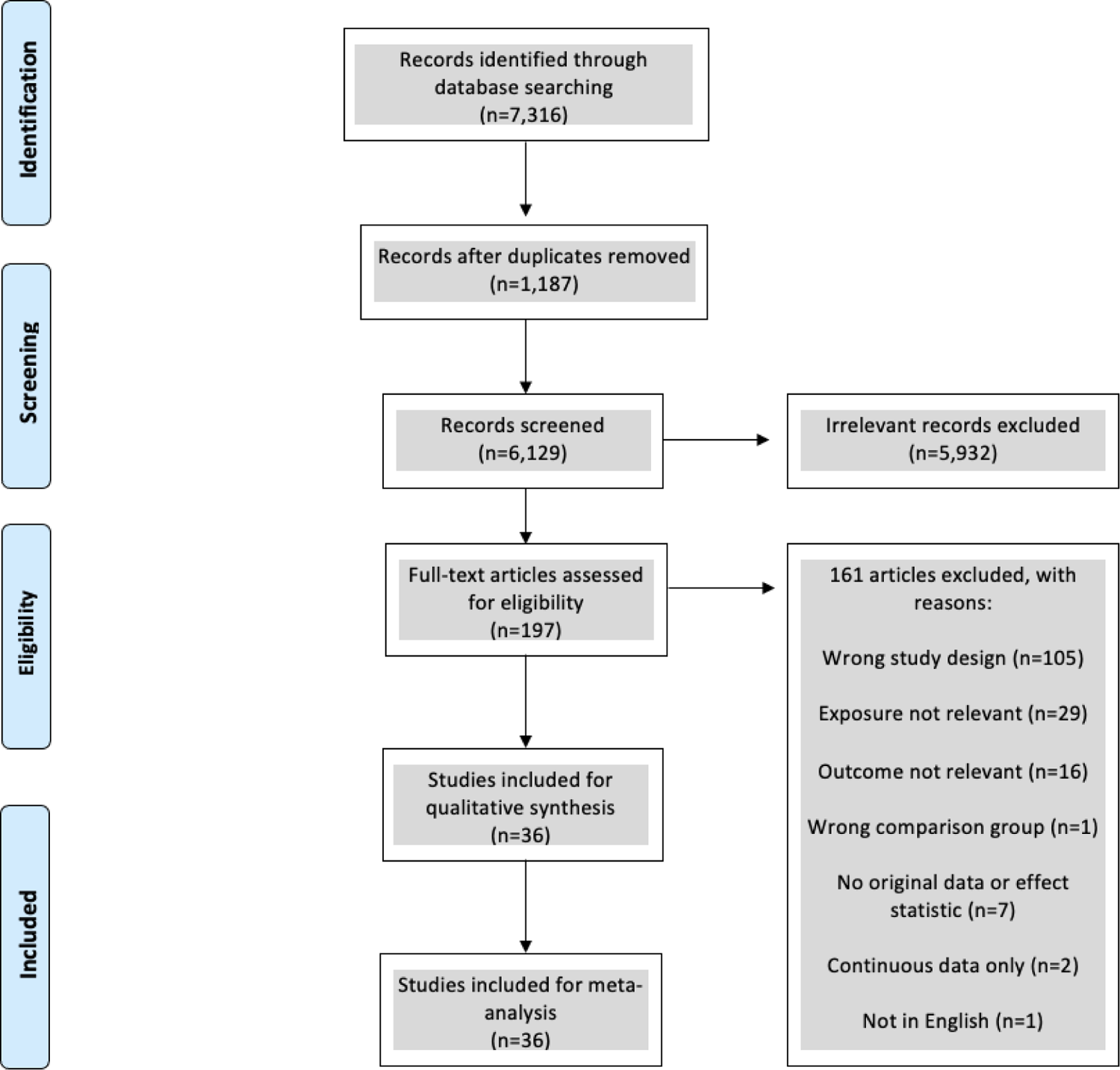
PRISMA flowchart representing the publication selection process.

This was performed independently by three authors (E.G., C.R and M.G.) using a standardised data collection spreadsheet containing: study ID; the first author’s last name, year of publication, study design, sample size, age and sex of the subjects, country of origin, duration of follow-up in cohort studies, assessment of exposure and outcome, adjusted covariates, number of cases, the effect estimates with 95%CIs and follow-up duration. From each study, risk estimates were extracted which reflected the greatest degree of adjustments for potential confounders. If study data appeared in more than one article, the most recent publication or largest e.g., pooled result were extracted for the statistical analysis.

### Exposure definitions

There are differences in metabolic trait definitions used, depending on the country where guidelines were written, age, sex, and ethnicity ^32–34^. For this study, the following metabolic traits definitions were used:

1. The WHO definition of obesity as body mass index (BMI) of ≥30 kg/m^2^ must have been used ^35^. BMI was acceptable if calculated from self-reported height and weight or taken from medical records or measured at baseline.
2. The WHO defines type 2 diabetes as a chronic disease that occurs when the body cannot effectively use the insulin it produces and is largely the result of excess body weight and physical inactivity. Diagnosis was defined as symptoms such as polyuria or polydipsia, plus^36,37^:

- random blood plasma glucose concentration ≥ 11.1 mmol/l or;
- fasting plasma glucose concentration ≥ 7.0 mmol/l (whole blood ≥ 6.1 mmol/l) or;
- two-hour plasma glucose concentration ≥ 11.1 mmol/l two hours after 75g anhydrous glucose in an oral glucose tolerance test (OGTT) or;
- haemoglobin (HbA1c) 6.5% or more (48 mmol/mol and above)

A diagnosis of type 2 diabetes via self-report, International Classification of Diseases (ICD) codes or medical records was also accepted.

1. The WHO definition of hypertension as ^38^:

- systolic blood pressure (SBP) reading of ≥130 mmHg; and/or
- diastolic blood pressure (DBP) readings on both days of ≥85 mmHg.

ICD-codes and patients taking antihypertensive medication were also acceptable diagnoses.

1. Dyslipidaemia classified as serum total cholesterol (TC), low density lipoprotein cholesterol (LDL-C), triglycerides, apolipoprotein B, or lipoprotein(a) concentrations above the 90^th^ percentile, or high density lipoprotein cholesterol (HDL-C) or apolipoprotein concentrations below the 10^th^ percentile for the general population in which the study was conducted ^39^. ICD-codes and patients taking cholesterol-lowering medication were also acceptable diagnoses.

### Outcome definitions

All diagnoses of incident head and neck squamous cell carcinoma should have been confirmed using histology by a trained pathologist and reported using appropriate ICD codes^40^. Cancer cases for the disease of interest have the following ICD-10 codes: oral cavity (C02.0 – C02.9, C03.0 – C03.9, C04.0 – C04.9, C05.0 – C06.9) oropharynx (C01.9, C02.4, C09.0 – C10.9), hypopharynx (C13.0 – C13.9), overlapping (C14 and combination of other sites) and 25 cases with unknown ICD code (other). Older versions of ICD code e.g., ICD-8 and 9 were also accepted.

### Quality assessment of studies

This assessment was used in evaluating the strength of evidence from the included studies. Three authors (A.G., C.R. and M.G.) independently assessed papers for risk of bias during data extraction using the Risk of Bias in Non-randomised Studies – of Exposures (ROBINS-E) tool (available at: https://www.riskofbias.info/welcome/robins-e-tool)^41^. The ROBINS-E tool aims to assess the result of interest in an observational study for risk of bias, where that study is designed to assess the effect of exposure on outcome ^42^. Pre-defined confounding variables that were agreed, included: age, sex, smoking status, alcohol intake, ethnicity, and socio-economic status. Further confounding variables that were recognised as important, but possibly to a lesser extent, included: dietary intake, physical exercise, and other metabolic traits (e.g., where BMI was the exposure, adjusting for hypertension, dyslipidaemia, and type 2 diabetes). The ‘appropriateness’ of each study to address the review question was also considered.

Data visualisations of the ROBINS-E tool were created using the *robvis* tool (available at: https://mcguinlu.shinyapps.io/robvis/). An individual panel has been developed for each individual exposure. Individual results from each included study were assessed using the tool for each included study. There was no difference in risk of bias assessment within each study and therefore the overall risk of bias assessment is presented below.

### Data analysis and synthesis

The analysis and synthesis process were performed in two phases. The first phase consisted of estimating the effect of each metabolic trait (obesity, type 2 diabetes, dyslipidaemia, and hypertension) on incident HNC separately for each included study. All included studies were cohort studies, with most using non-exposed comparisons and reporting risk (RR) and hazard ratios (HR). Other studies using external general population comparisons reported standardised incidence ratios (SIR). Because the absolute risk of HNC is low, all the above measures yield similar estimates of RR and this approach has been taken previously in similar head and neck cancer meta-analyses ^17^. The second phase focussed on estimating the combined effect across studies using meta-analysis, using *metan* software in Stata (*Stata Statistical Software: Release 16*. College Station, TX: StataCorp LLC). Summary effects with their corresponding 95%CIs were derived using the method described by DerSimonian and Laird for both fixed effects and random effects models, which incorporates between-study variability ^43^. This was performed separately for each metabolic trait, with a preference for random-effects if there is no important difference between the two methods.

The analysis yielded information about the heterogeneity of effects across studies, using Cochran’s Q test and Higgins’ I^2^ statistic. To further explore the sources of heterogeneity, subgroup, and meta-regression analyses were performed according to age, geographic location, year of publication, methods of exposure assessment, length of follow-up, the number of cancer cases and adjustment for confounding factors including age, sex, smoking and alcohol use. Small study effects, which may reflect publication bias, study quality or other sources of heterogeneity were assessed using funnel plots, which show effect estimates from individual studies plotted against the standard error as a measure of each study’s size or precision. In the absence of bias and between study heterogeneity, the scatter will be due to sampling variation alone, with the plot resembling a symmetrical inverted funnel. Smaller studies will be scattered more widely at the bottom, with larger studies nearer the top and any gaps where smaller studies are missing will be evident ^44^.

Sensitivity analyses were performed by excluding two large pooled analyses from the meta-analyses, one for BMI ^11^ and the other for type 2 diabetes ^17^, and calculating an estimate for the remainder of the studies to evaluate whether the results were significantly affected by these studies.

## Results

The search generated 7,316 potential records of which 6,129 were screened following the removal of 1,187 duplicates. Of these, 197 full-text articles of interest were assessed for eligibility. One hundred and sixty one of these articles were subsequently excluded: 105 due to study designs that could not infer a temporal relationship between the exposure and outcome, 29 where the exposure was not relevant or as defined above, 16 in which the outcome was not relevant or as defined above, 1 with an ineligible comparison group (diabetics treated versus not treated), 7 with no original data or reported effect statistic (e.g., short abstracts), 2 studies had continuous data only which was not comparable and 1 article that was not written in English. The remaining 36 eligible articles were included for full qualitative and quantitative synthesis to investigate the association between metabolic traits and HNC risk (Figure 1). One large pooled analysis for BMI ^11^ included the results from 4 of the studies ^45–48^ and similarly, a pooled analysis for type 2 diabetes^17^ included the results from 6 of the studies ^9,49–53^, so only the two pooled analyses were represented in the final **Table 2** and subsequent analyses.

**Table 2.**
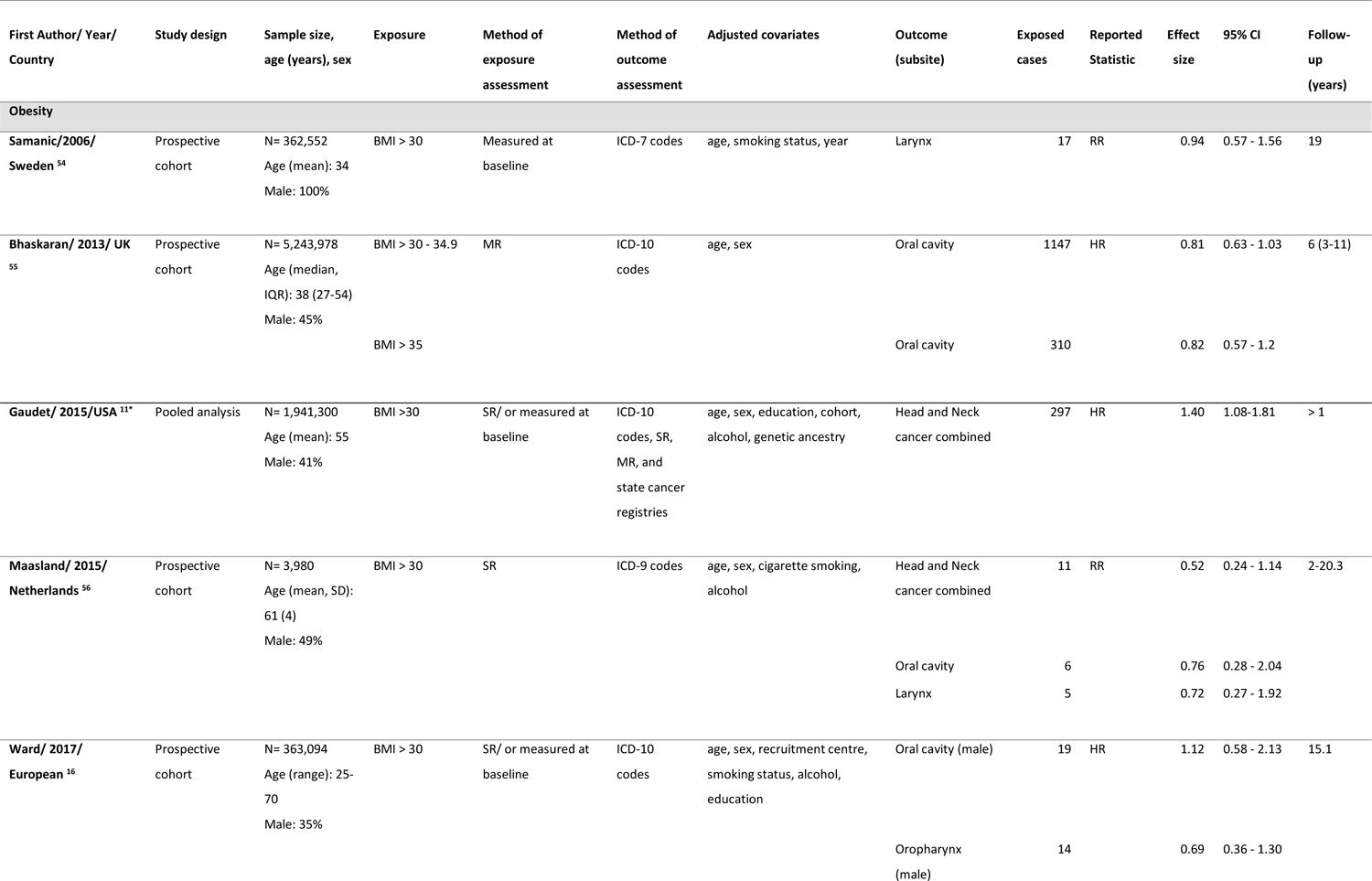
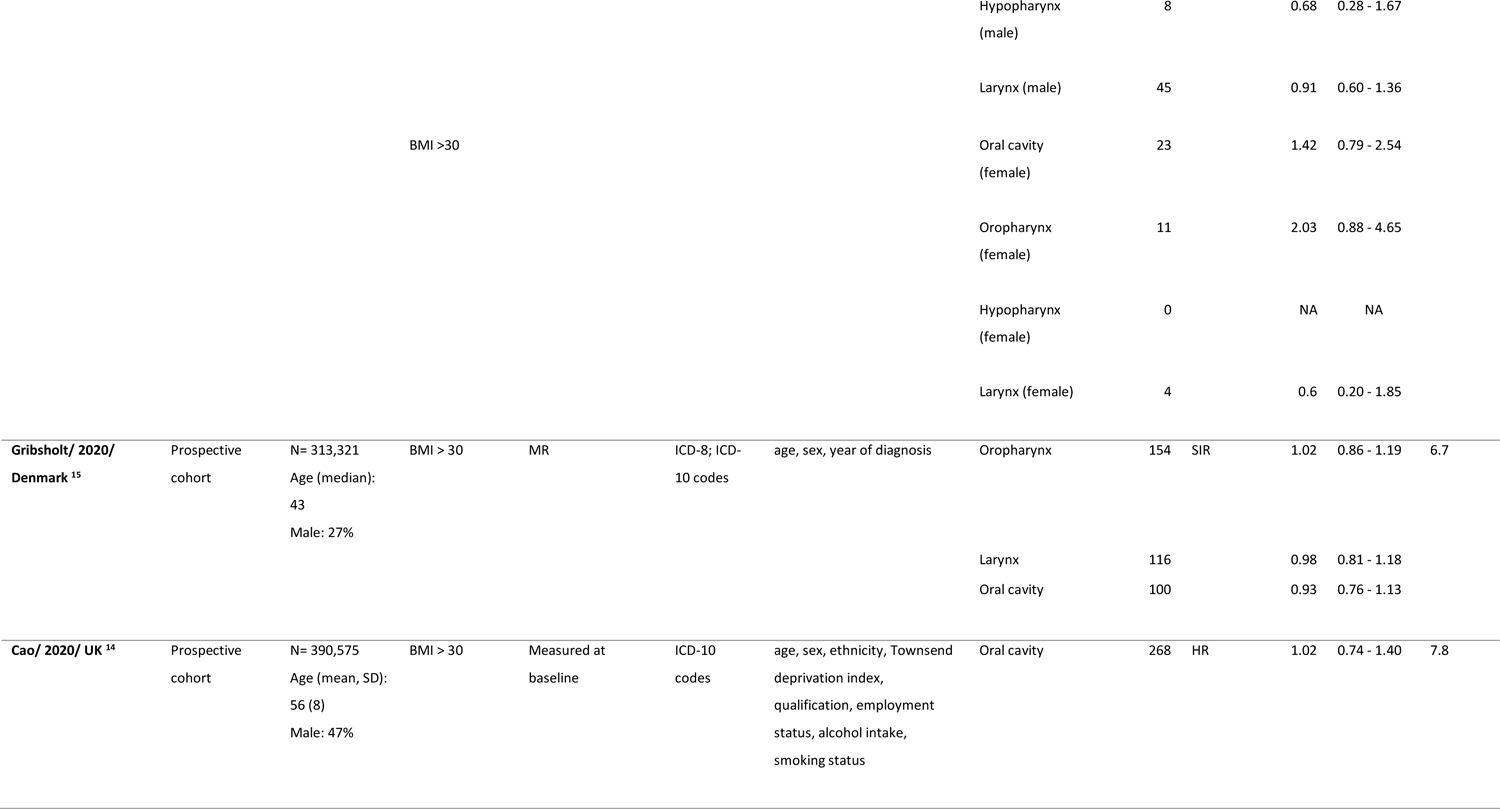
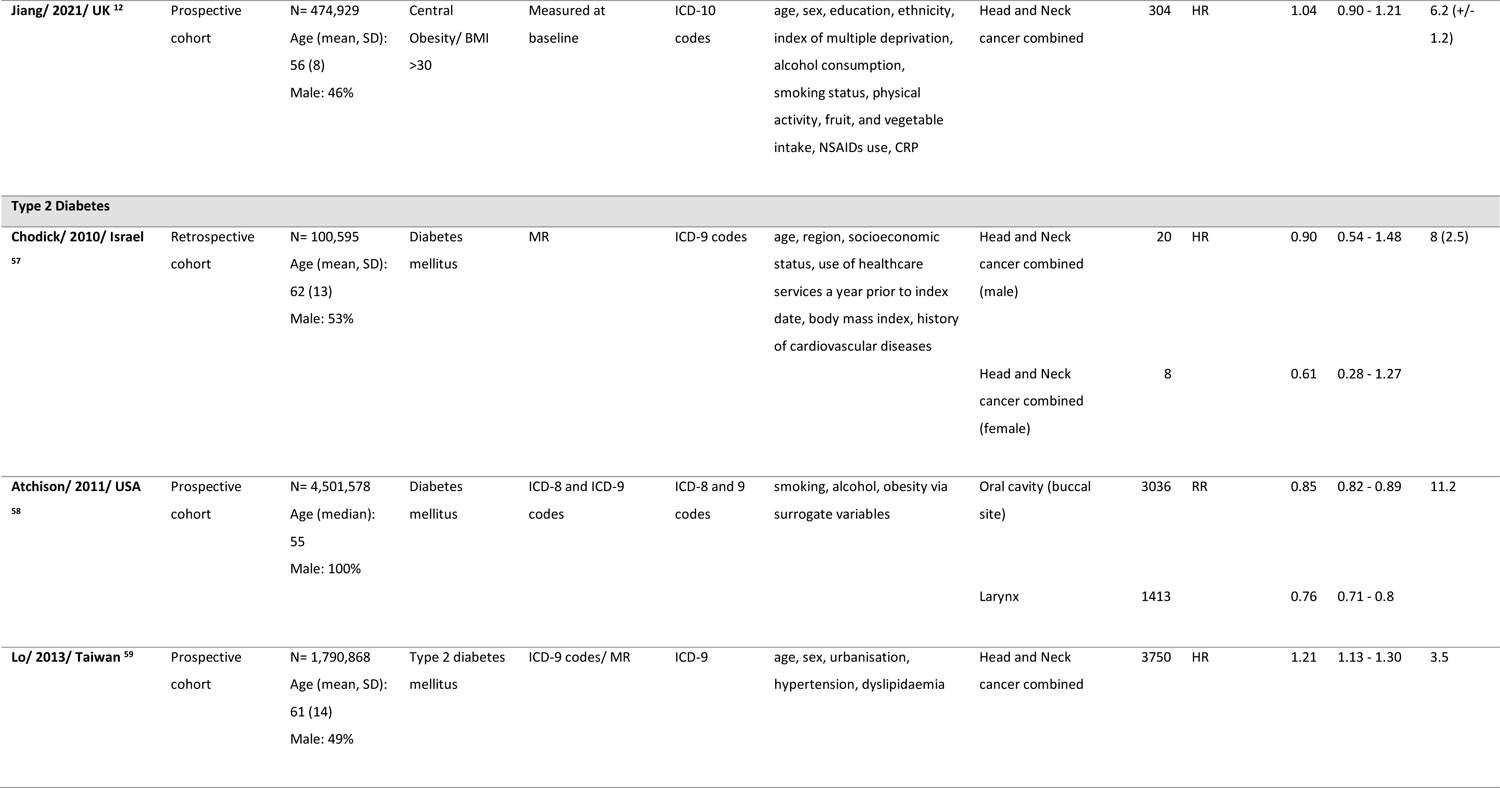
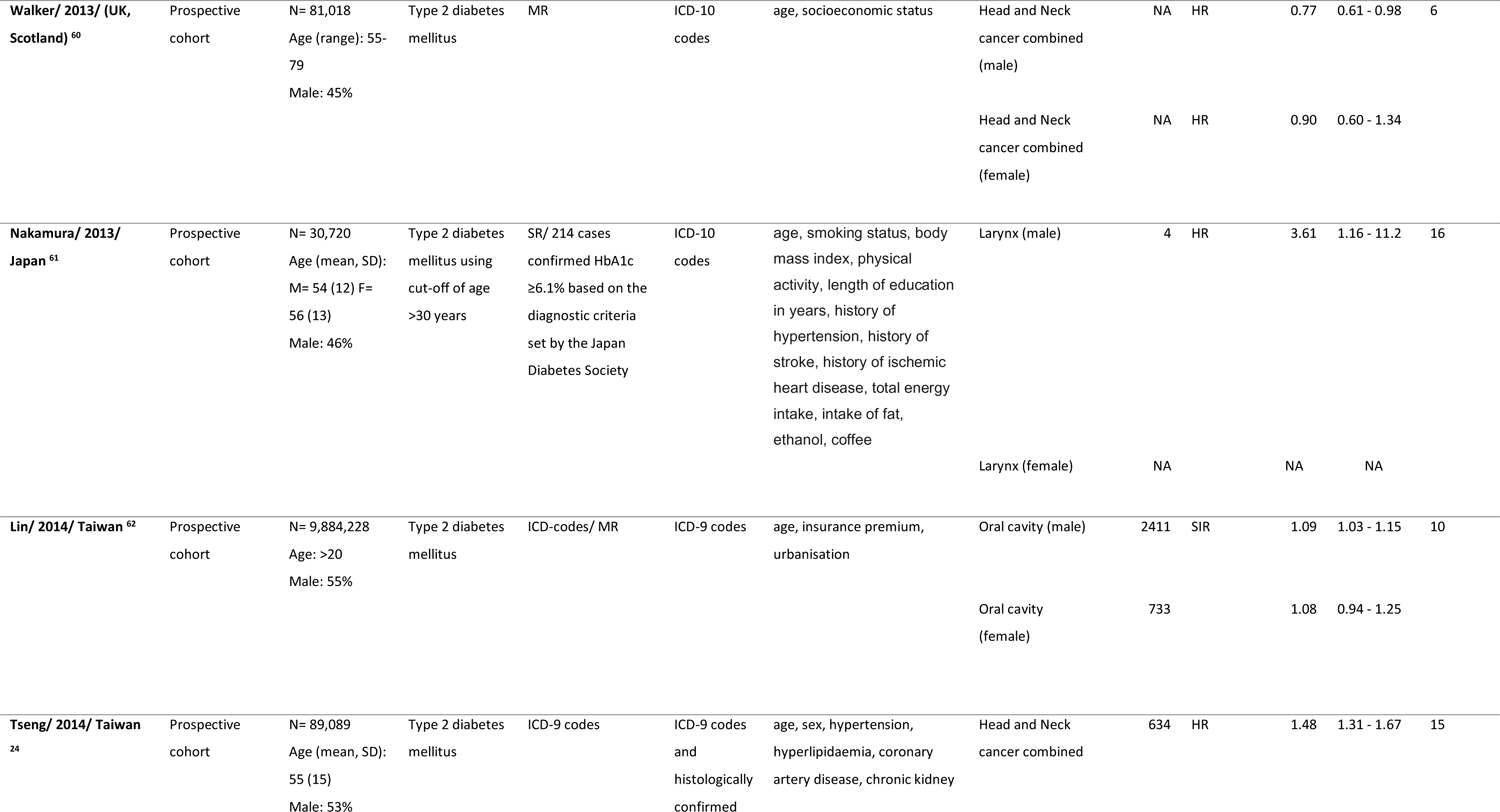
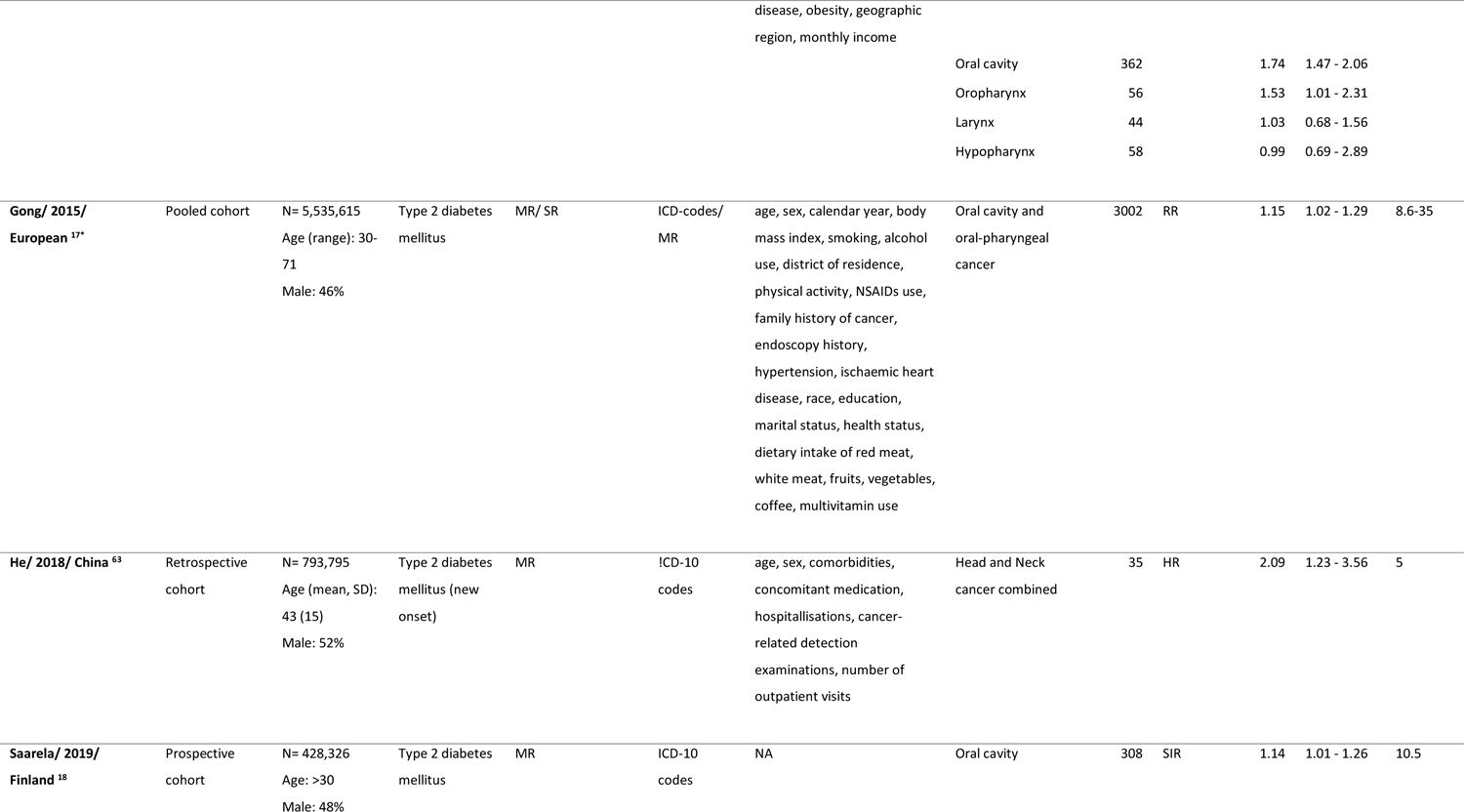
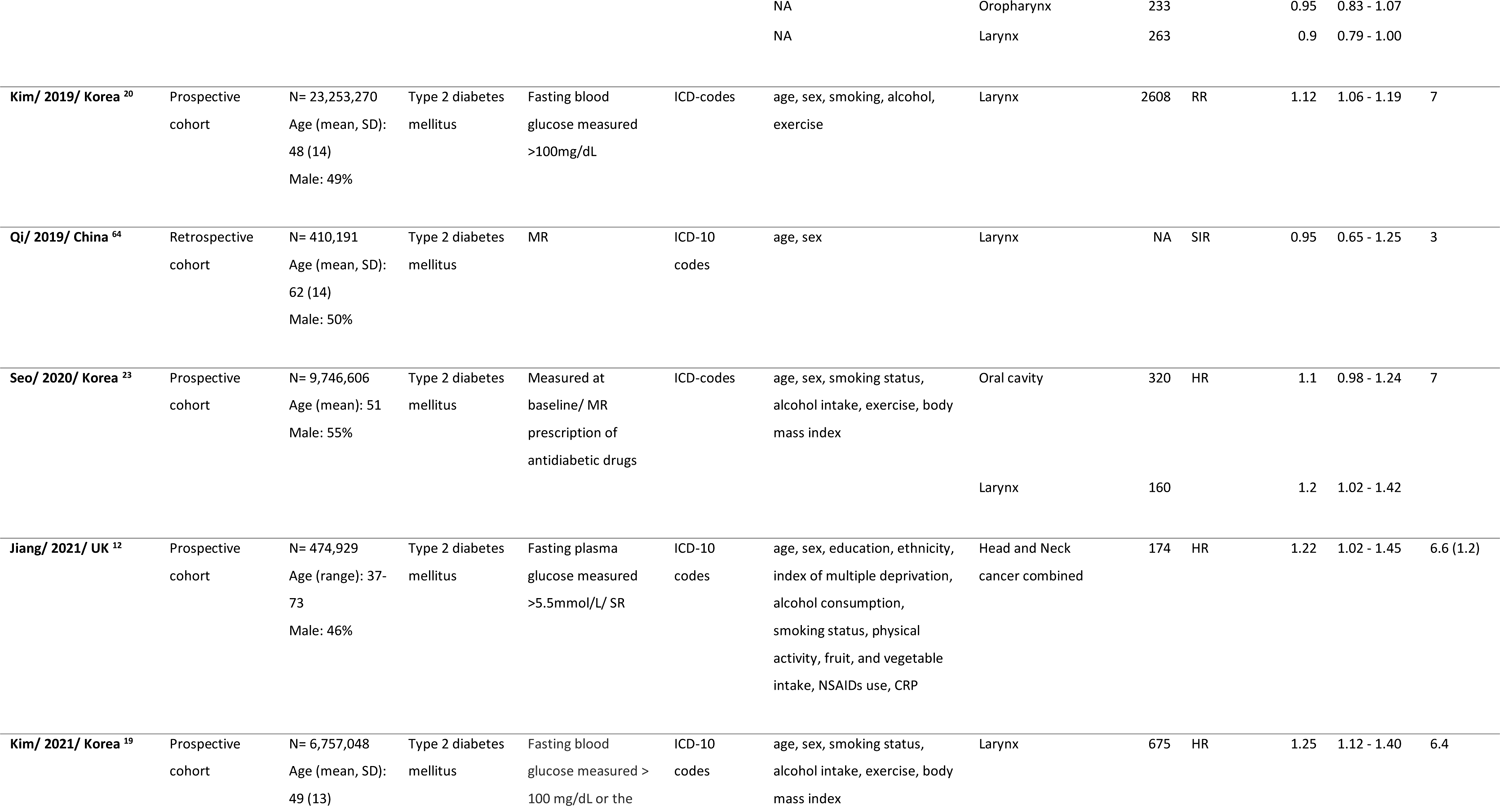
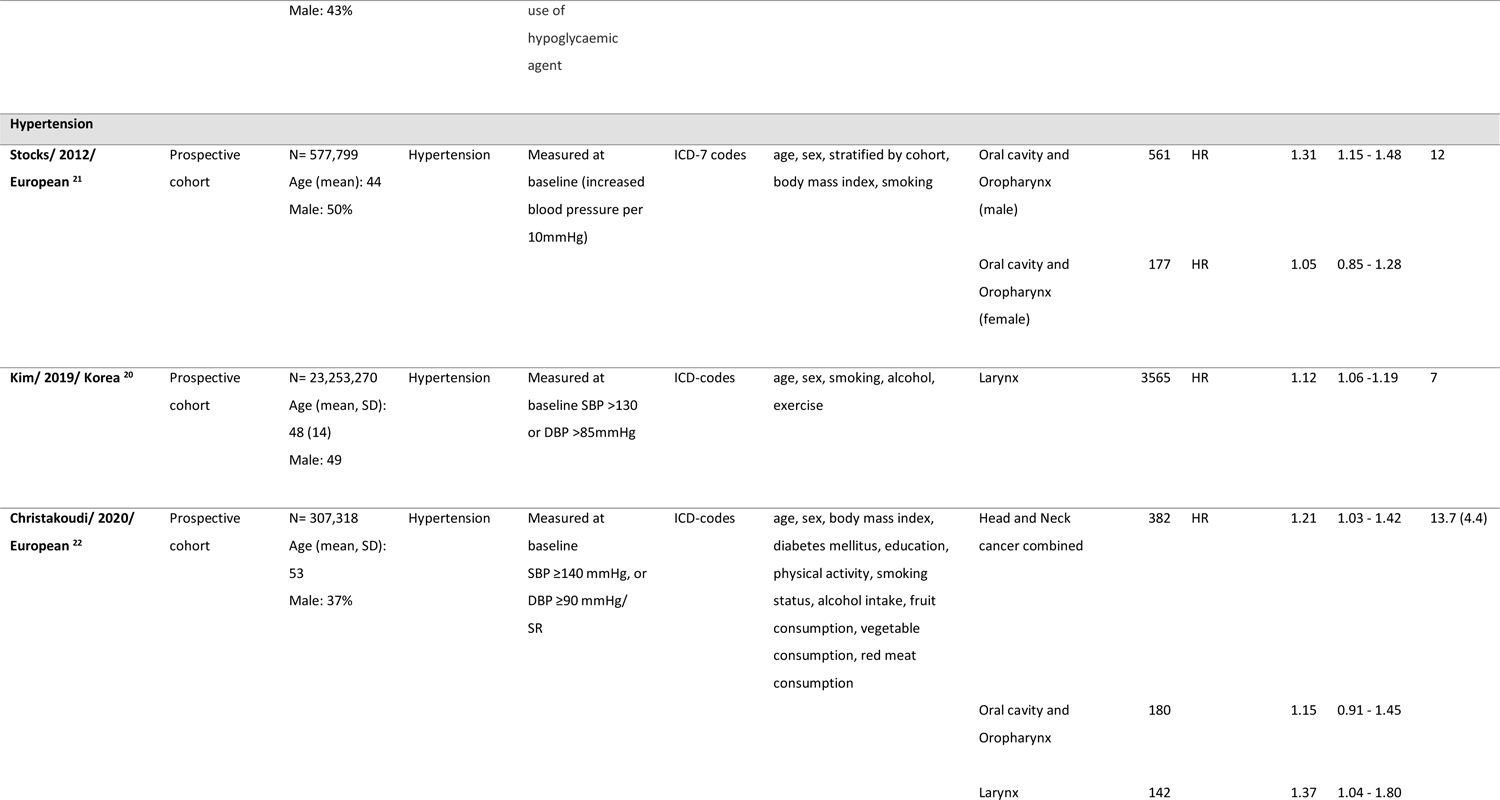
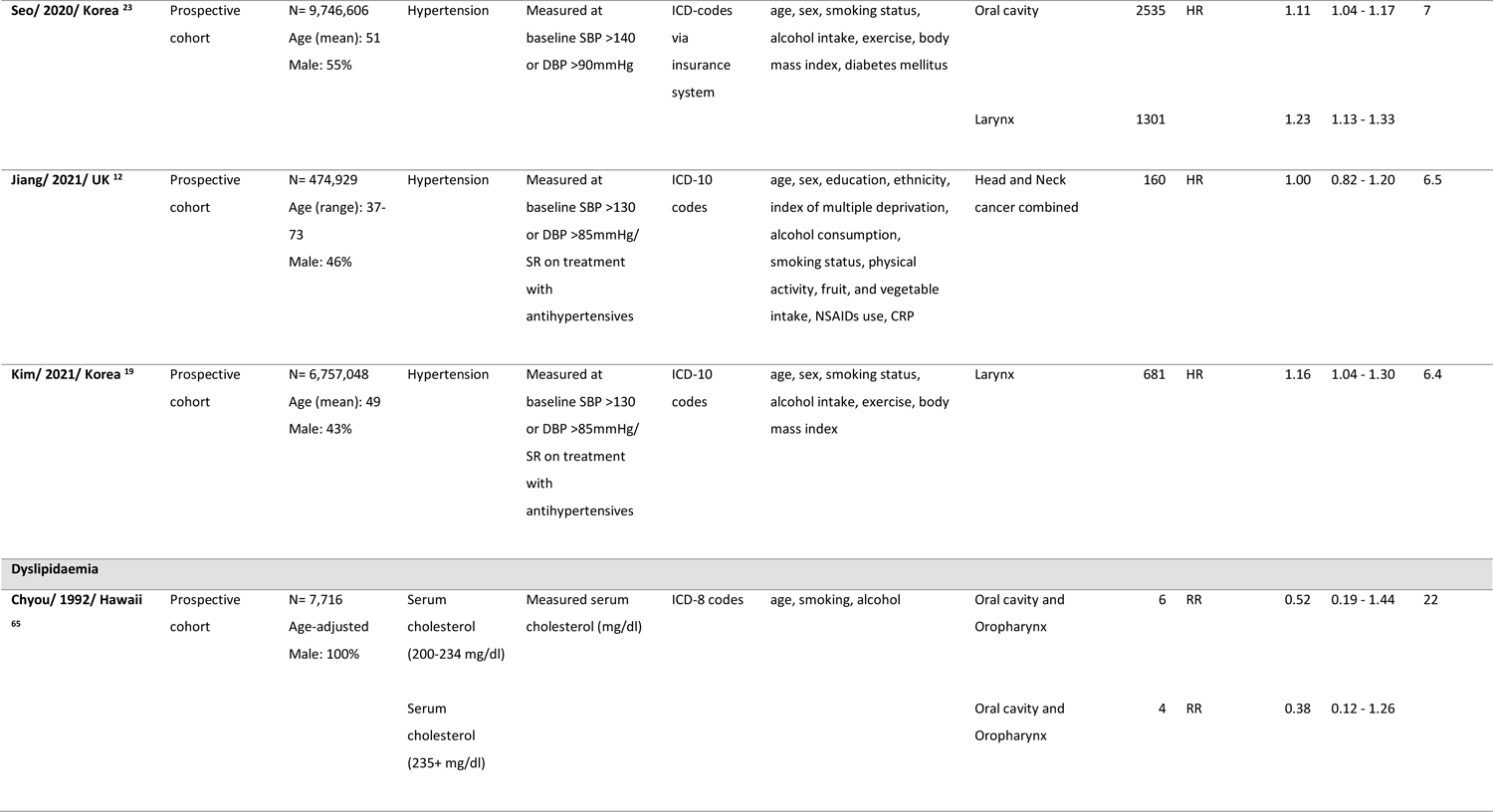
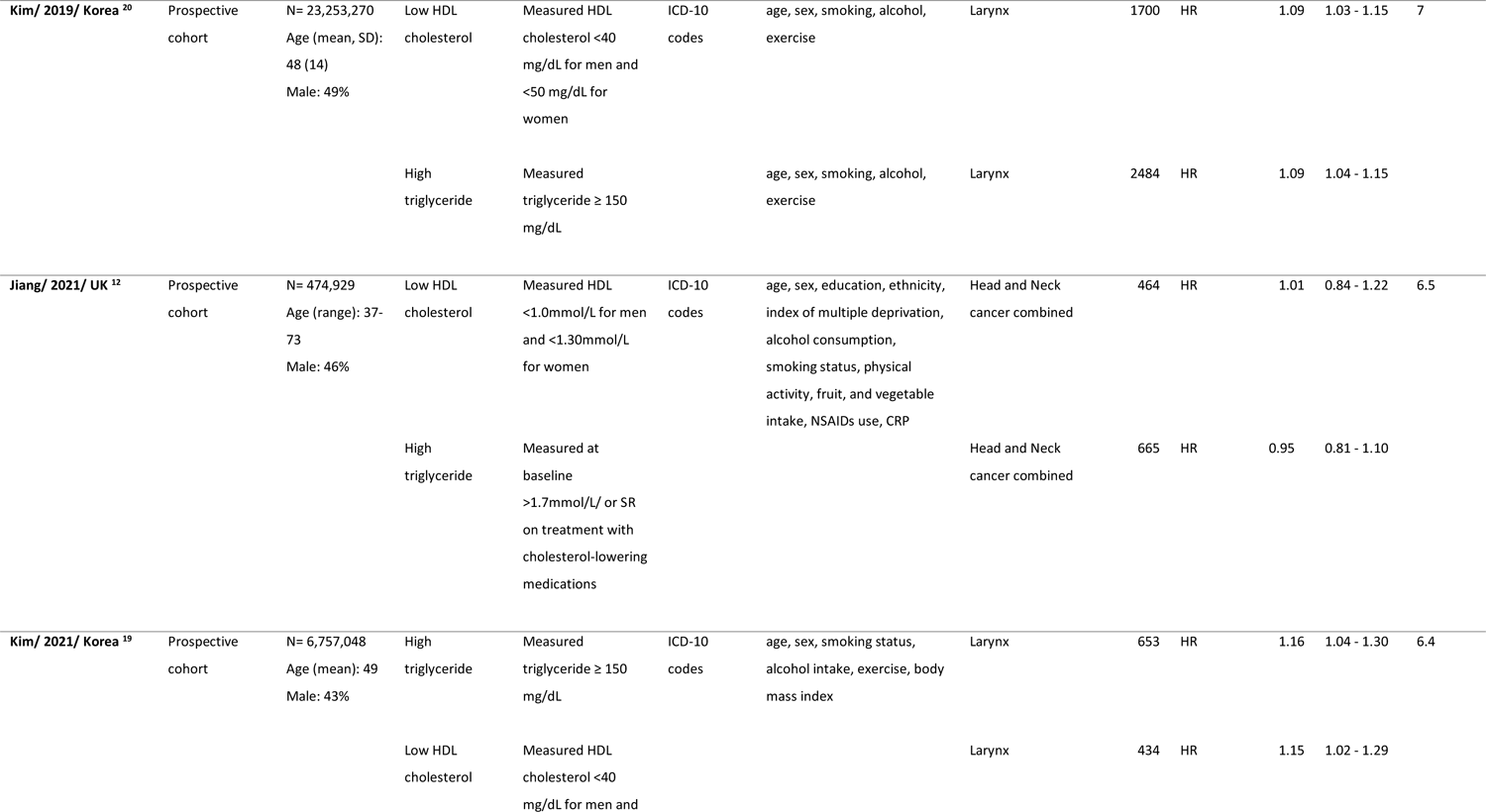
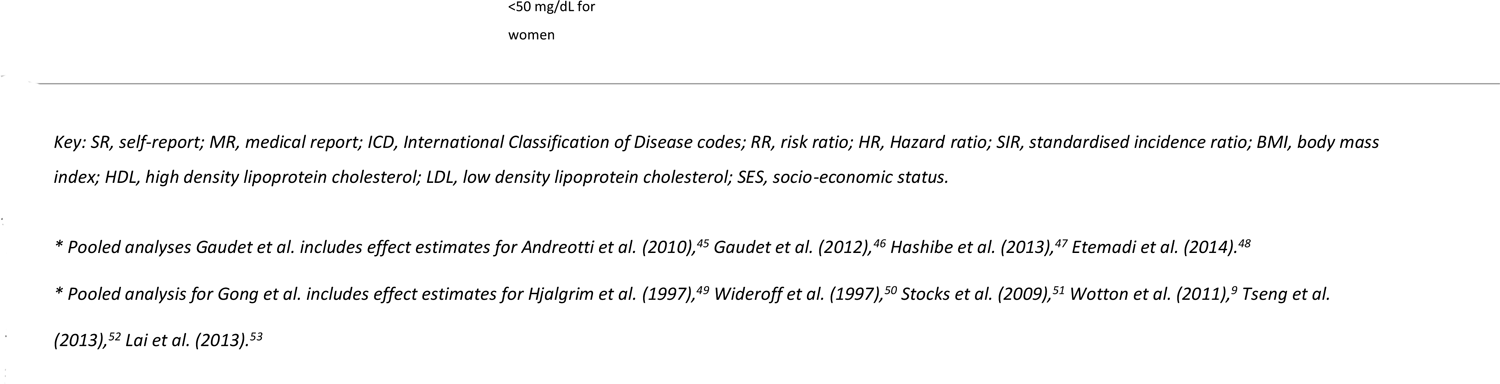
Descriptive characteristics of studies of metabolic traits and risk of head and neck c3a2n8cer.

Eight studies reported the association between BMI (n= 2,859 cases) and risk of HNC ^11,12,14–16,54–56^, with 15 studies reporting the association with type 2 diabetes (n= 17,582 cases) ^12,17–20,23,24,57–64^, six studies hypertension (n= 12,151 cases) ^12,19–23^, and four studies dyslipidaemia (n= 6,410 cases) ^12,19,20,65^ and HNC risk (**Table 2**). Adjustments were made for potential confounders of two or more factors, including age, in all studies.

### Obesity and risk of HNC

In the analysis of five studies investigating the association between obesity and incidence of combined head and neck cancer sites, there was an overall RR of 1.06 (95%CI (0.76, 1.49)) using a random-effects model, little evidence of an effect (Figure 2a). The result was similar when using a fixed-effects model (RR= 1.10, 95%CI (0.97, 1.25)). There was however evidence for the presence of substantial heterogeneity between studies (*P* heterogeneity <0.024, I^2^= 73.2%) (Figure 2a). With respect to individual subsites, there was again little evidence of an association of obesity on oral cavity (RR= 0.92, 95%CI (0.81, 1.04)), oropharyngeal (RR= 1.05, 95%CI (0.69, 1.59)), or laryngeal (RR= 0.95, 95%CI (0.81, 1.11)) cancer using a random-effects model. There were similar results when using a fixed-effects model (Figure 2b – d).

**Figure 2.**
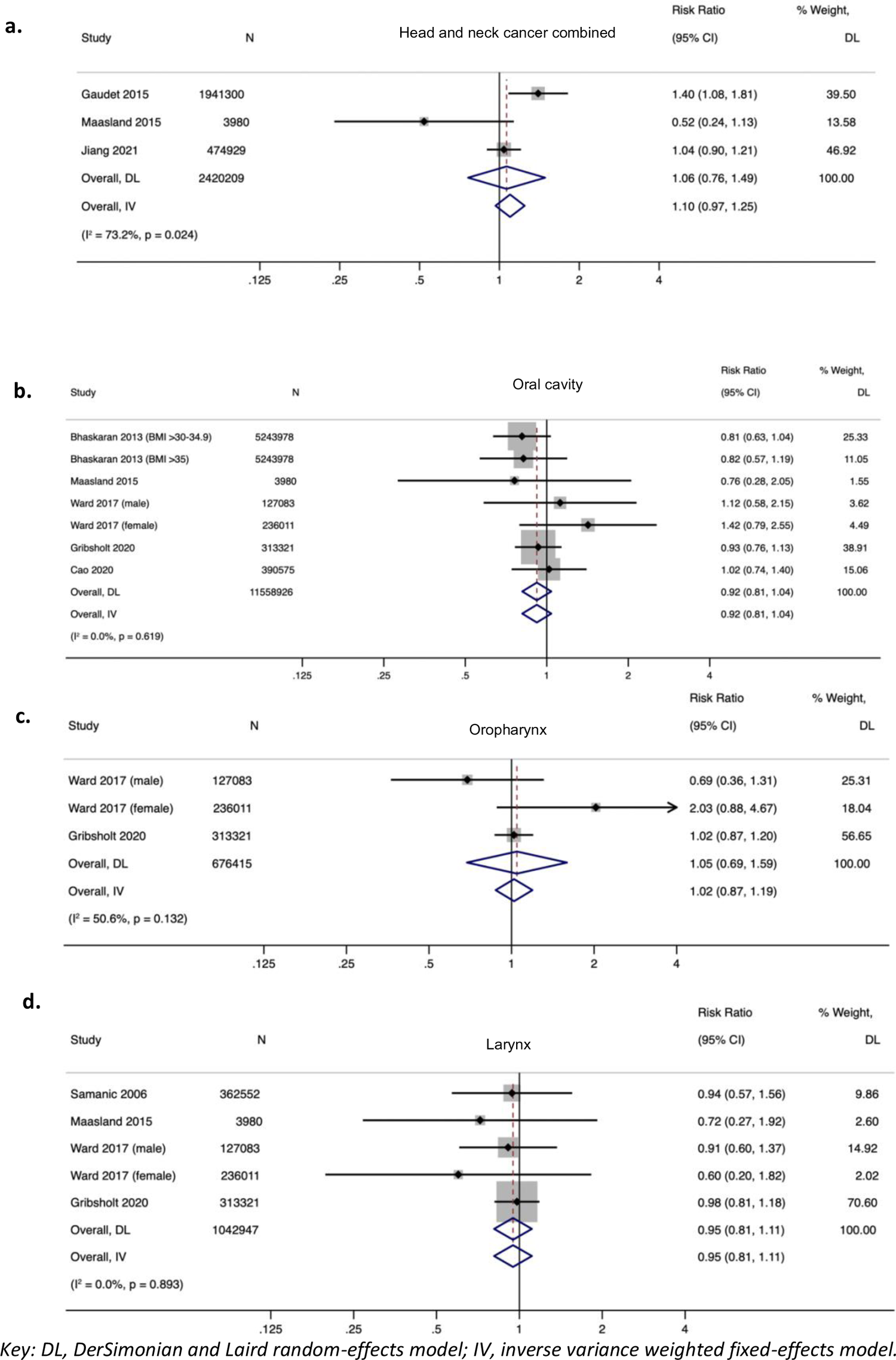
Relative risks for the association between obesity and incidence of head and neck cancer combined and by subsite.

### Type 2 diabetes mellitus and risk of HNC

In the analysis of 6 studies on the association between type 2 diabetes and incidence of combined head and neck cancer, there was an overall RR of 1.13 (95%CI (0.95, 1.34)) using a random-effects model. Strong evidence of heterogeneity was present between studies (*P* heterogeneity= <0.0001, I^2^= 80.0%) (Figure 3a). There was a trend towards an increased risk association with oral cavity (RR= 1.13, 95%CI (0.97, 1.31)), oropharyngeal (RR= 1.16, 95%CI (0.73, 1.83)) and laryngeal (RR= 1.05, 95%CI (0.87, 1.26)) cancer using a random effects model, but again all results crossed the null, with strong evidence of heterogeneity across all subsites (Figure 3b – d). When using a fixed effects model, the result for combined head and neck cancer sites showed a stronger increased risk effect (RR= 1.22, 95%CI (1.16, 1.29)), but attenuated towards the null in the individual subsites.

**Figure 3.**
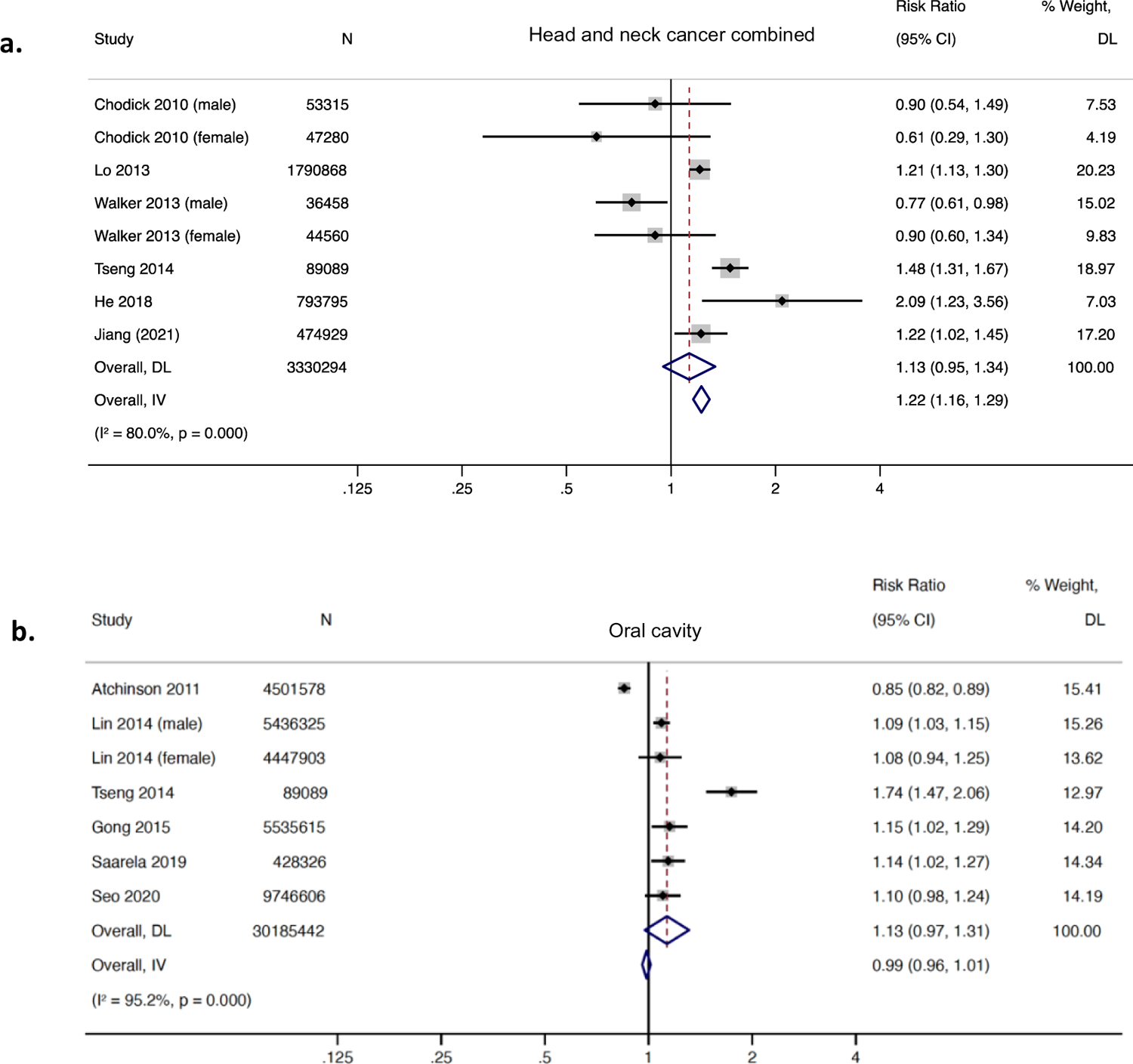
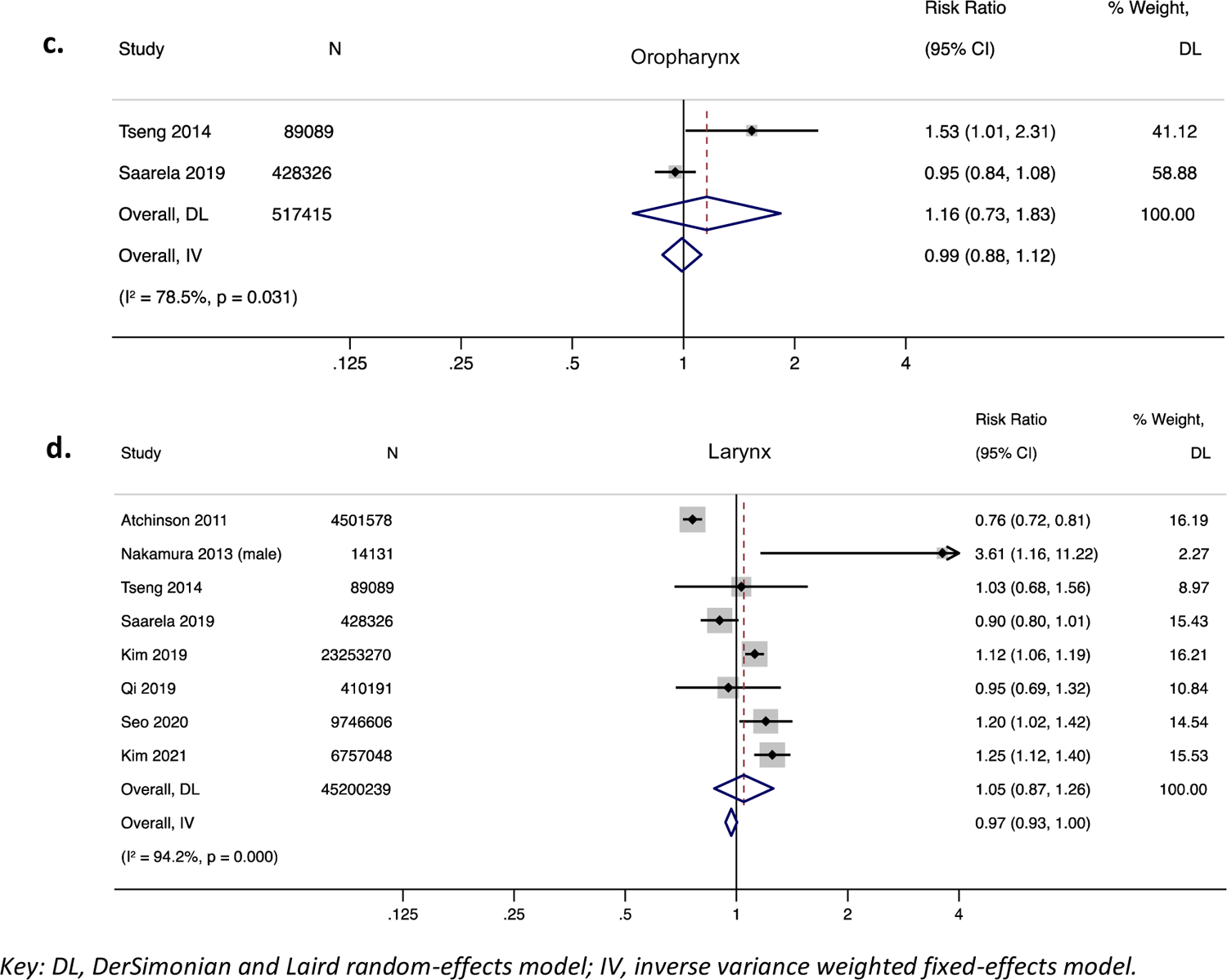
Relative risks for the association between type 2 diabetes mellitus and incidence of head and neck cancer combined and by subsite.

### Hypertension and risk of HNC

In the analysis of only 2 studies on the association between hypertension and incidence of combined head and neck cancer, there was limited evidence of an increased risk (RR= 1.11, 95%CI (0.92, 1.33)) using a random-effects model. There was evidence of substantial heterogeneity among studies (*P* heterogeneity= 0.134, I^2^= 55.6%) (Figure 4a). An increased risk of hypertension was consistent across HNC subsites, with a RR of 1.16 (95%CI (1.05, 1.28)) found for combined oral cavity and oropharyngeal cancer, and the strongest effect found in the larynx (RR= 1.17, 95%CI (1.10, 1.25)) (Figure 4b – c). The amount of heterogeneity was similar in both oral cavity and oropharyngeal combined (*P* heterogeneity= 0.108, I^2^= 50.6%) and laryngeal cancer analyses (*P* heterogeneity= 0.186, I^2^= 37.7%). The results remained similar when using a fixed effects model across all sites.

**Figure 4.**
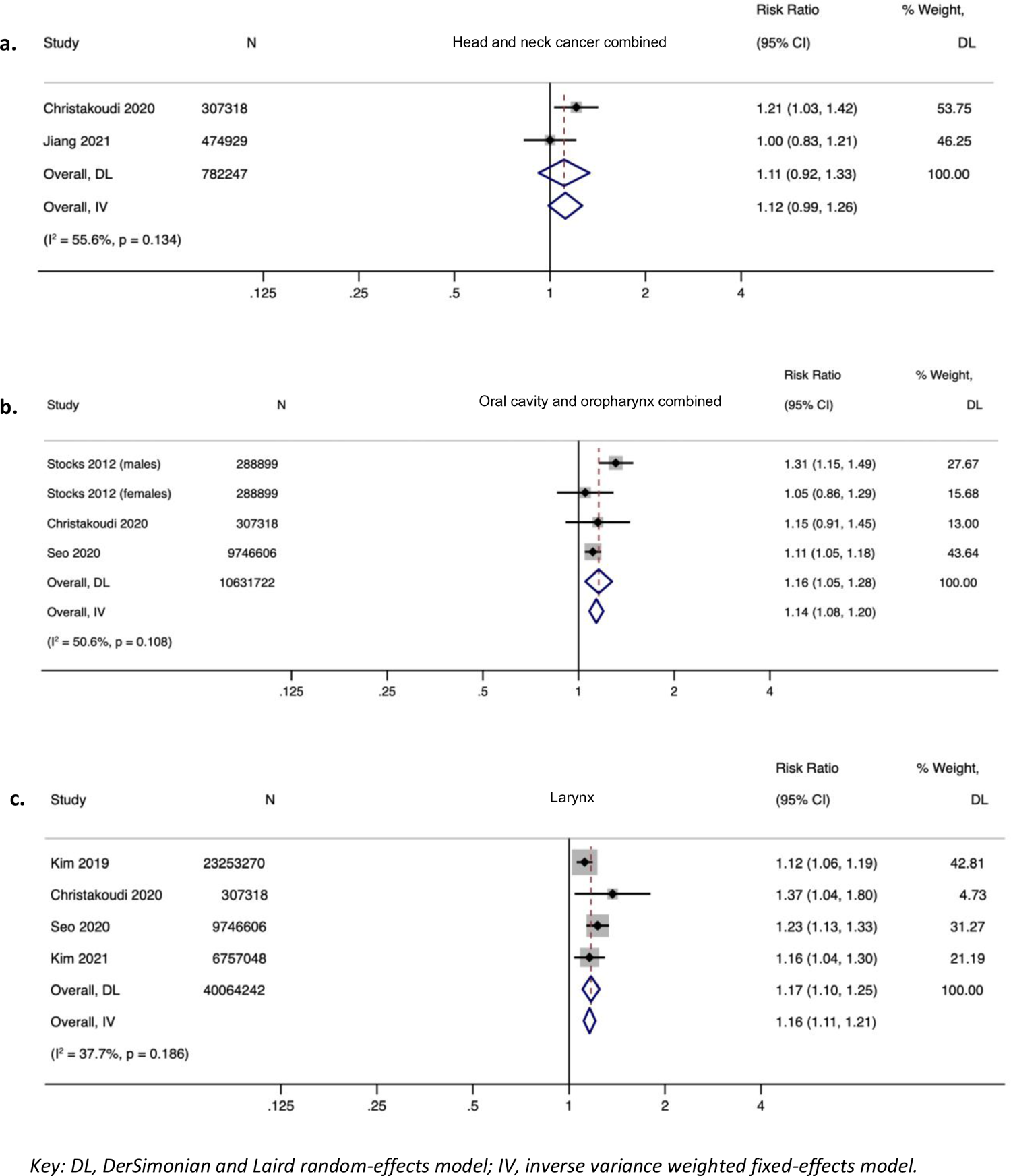
Relative risks for the association between hypertension and incidence of head and neck cancer combined and by subsite.

### Dyslipidaemia and risk of HNC

There were only 2 studies available for meta-analysis, both investigating the effect of dyslipidaemia on incident laryngeal cancer risk. There was some evidence of an increased risk association of both low HDL (RR= 1.12, 95%CI (1.07, 1.18)) and high triglyceride levels (RR= 1.10, 95%CI (1.05, 1.15)) using a random-effects model (Error! Not a valid bookmark self-reference.**a – b**). In the low HDL analysis, there was evidence of moderate heterogeneity (*P* heterogeneity= 0.103, I^2^= 62.5%), which was not found in the high triglycerides analysis (*P* heterogeneity= 0.319, I^2^= 0.0%) (Error! Not a valid bookmark self-reference.**a – b**). The results remained similar when using a fixed effects model across all sites. These findings were not supported in the single study by Jiang *et al.*^12^ which investigated low HDL (RR= 1.01, 95%CI (0.84, 1.22)) and high triglycerides (RR= 0.95, 95%CI (0.81, 1.10)) on head and neck combined cancer (**Table 2**).

### Subgroup analyses

Pre-defined subgroup meta-analyses were conducted for all four metabolic traits according to age, geographic location, year of publication, methods of exposure assessment, risk of bias score, length of follow-up, the number of cancer cases and adjustment for confounding factors including age, sex, smoking and alcohol use (**Supplementary Table 3**).

Stratifying by geographic location of the study resulted in a difference for both obesity (European (RR= 0.97, 95%CI (0.90, 1.04)) vs USA (RR= 1.40, 95%CI (1.08, 1.81), *P* difference= 0.007), type 2 diabetes (Asian (RR= 1.23, 95%CI (1.13, 1.33)) vs European (RR= 1.00, 95%CI (0.90, 1.12)) vs USA (RR= 0.81, 95%CI (0.72, 0.90), *P* difference <0.0001) and dyslipidaemia (Asian (RR= 1.12, 95%CI (1.08, 1.16) vs European (RR= 0.97, 95%CI (0.87, 1.10) vs USA (RR= 0.45, 95%CI (0.21, 0.98), *P* difference= 0.006). Various methods of exposure assessment did result in a differences for both obesity and type 2 diabetes. Within obesity, there were differences between the use of medical records (RR= 0.94, 95%CI (0.86, 1.04) vs self-report (RR= 0.63, 95%CI (0.38, 1.06)) vs measurements taken at baseline (RR= 1.02, 95%CI (0.91, 1.15)) vs a combination of the aforementioned methods (RR= 1.40, 95%CI (1.08, 1.81)) (*P* difference = 0.011).

When stratifying by year of publication (before or after 2010), by adjustment for confounding factors, length of follow-up and number of cancer cases, this resulted in differences only within dyslipidaemia. However, there was only one small study by Chyou *et al.* (1992)^65^ representing the before 2010, no adjustment, >10 year follow-up and <50 cancer cases categories which is of high risk of bias.

### Risk of bias assessment

Risk of bias assessment was undertaken using the Risk of Bias In Non-randomized Studies – of Exposure (ROBINS-E) tool ^42^. From all included studies, each result (n= 85) included in the meta-analysis was individually risk assessed across the seven domains of bias, accounting for confounding variables which were pre-defined as important (i.e., age, sex, smoking status, alcohol intake, ethnicity, and socio-economic status). There was no difference between individual results within each study, therefore the overall study ratings only are presented in **Supplementary Figures 1 – 4**. One study was rated as ‘Very high’ risk of bias (3.7%) ^18^, 21 studies were rated as ‘High’ (77.8%) ^15–17,19–24,54–65^, one study as ‘Some concerns’ (3.7%) ^66^ and the remaining studies as ‘Low’ (14.8%) ^12,14^. All ‘High’ and ‘Very high’ ratings were due to bias that arose due to inadequate adjustment for confounding and/or the handling of missing data.

Funnel plots for each metabolic trait were used to assess asymmetry which may be as a result of true heterogeneity, poor methodological quality in smaller studies, reporting or publication bias ^44^. For obesity, there appeared to be only limited asymmetry, with only one outlier by Gaudet *et al.*^11^ and potential smaller studies missing from the bottom right corner (Figure 6a). For type 2 diabetes mellitus, there were significantly more larger studies, many of which lay outside the 95%CI suggesting the presence of heterogeneity. There were eight results from several studies which appear to be outliers in the type 2 diabetes funnel plot, including results from Atchison *et al.* ^58^, Lo *et al.* ^59^, Tseng *et al.* ^24^, Lin *et al.* ^62^, and Kim *et al.* ^19,20^. Four out of five of these outlier studies were distinct in that they were conducted in Asian only populations (Figure 6b). There were too few (≤10) studies for hypertension and dyslipidaemia) to confidently distinguish true asymmetry (Figure 6c-d).

### Sensitivity analyses

No association was found when dropping the largest pooled analysis by Gaudet *et al.*^11^ for obesity in combined head and neck cancer (RR= 0.82, 95%CI (0.43, 1.56) with evidence again for heterogeneity (I^2^ = 65.9%, *P heterogeneity* = 0.087). This lack of effect for obesity was consistent across subsites, with results all crossing the null. Similarly, there was no difference in the effect of type 2 diabetes on oral cancer risk when leaving out the other large pooled analysis by Gong *et al.*^17^ (RR= 0.98, 95%CI (0.95, 1.01), with no change in evidence for substantial heterogeneity (I^2^ = 95.8%, *P heterogeneity =* <0.001).

## Discussion

This is the most comprehensive and up-to-date systematic review and meta-analysis on this topic, considering the association of metabolic traits on HNC across subsites at a global population level. Despite observations from individual studies suggesting an association between BMI and HNC, no effect was found within any subsite in this study, using random-effects meta-analyses (Figure 2). There was also limited evidence of an association for type 2 diabetes (Figure 3), but perhaps a trend towards increased risk for overall HNC, which is in keeping with previous pooled estimates ^12,19,20,59^. Conversely, there was evidence of an increased HNC risk for both hypertension and dyslipidaemia across subsites, but with fewer studies included in these meta-analyses (Figure 4 & Figure 5). Furthermore, over 80% of studies were judged to be at ‘Very High’ or ‘High’ risk of bias using the ROBINS-E tool (**Supplementary Figures 1 – 4**), so caution must be exercised when interpreting these results. Of those studies assessed as low risk of bias, their results suggest limited evidence for an effect of obesity, but an increased risk association of type 2 diabetes ^12^ on HNC risk. Conversely, this was not the case for hypertension and dyslipidaemia, where the one study assessed as low risk of bias for these traits did not provide evidence of an effect on HNC risk ^12^. Variability in the individual study results may exist due to differences in timing of exposure measurement, confounder adjustment, or from other sources of bias.

**Figure 5.**
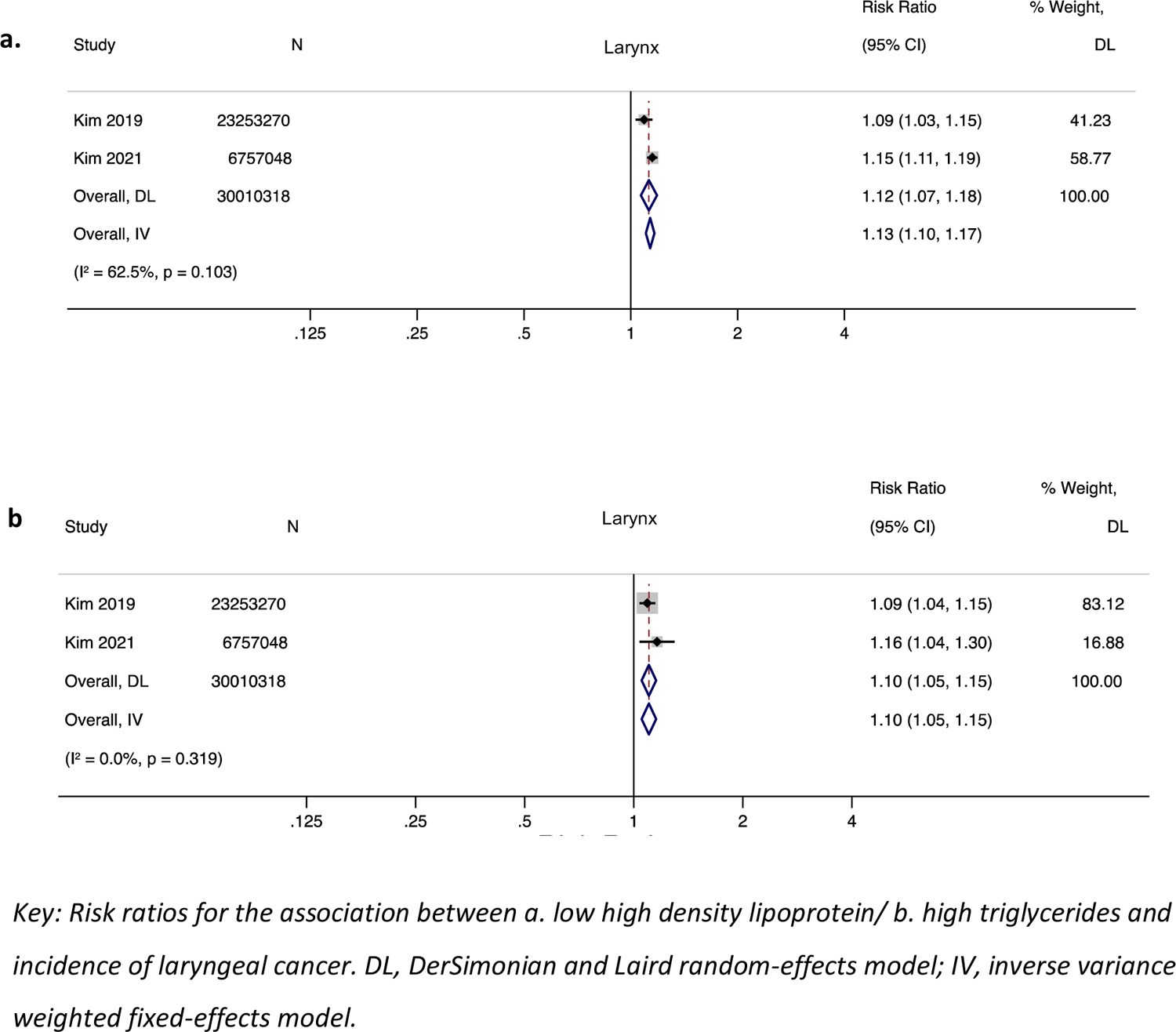
Relative risks for the association between dyslipidaemia and incidence of laryngeal cancer.

**Figure 6.**
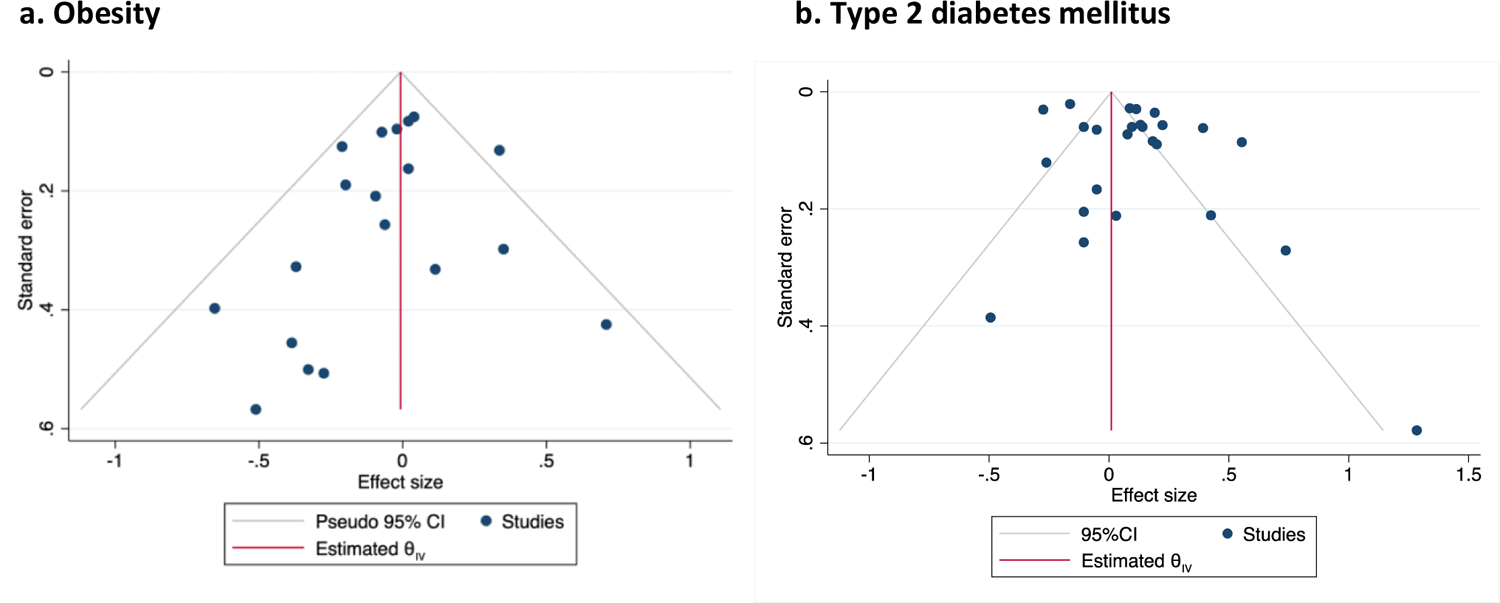
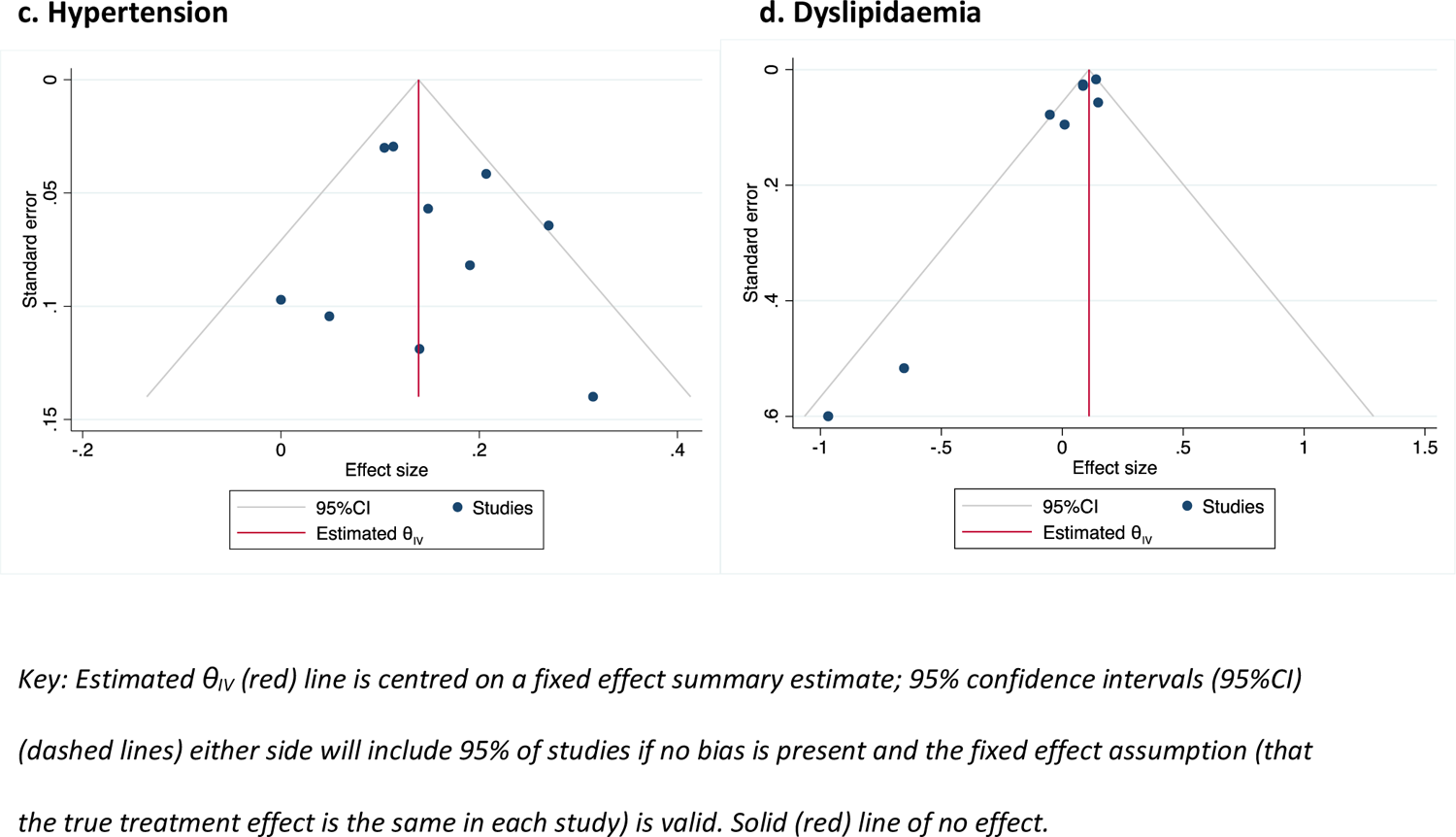
Funnel plots showing effect estimates for metabolic traits on overall head and neck cancer isk (all subsites)

### Dyslipidaemia and risk of HNC

There were only 2 studies available for meta-analysis, both investigating the effect of dyslipidaemia on incident laryngeal cancer risk. There was some evidence of an increased risk association of both low HDL (RR= 1.12, 95%CI (1.07, 1.18)) and high triglyceride levels (RR= 1.10, 95%CI (1.05, 1.15)) using a random-effects model (Error! Not a valid bookmark self-reference.**a – b**). In the low HDL analysis, there was evidence of moderate heterogeneity (*P* heterogeneity= 0.103, I^2^= 62.5%), which was not found in the high triglycerides analysis (*P* heterogeneity= 0.319, I^2^= 0.0%) (Error! Not a valid bookmark self-reference.**a – b**). The results remained similar when using a fixed effects model across all sites. These findings were not supported in the single study by Jiang *et al.*^12^ which investigated low HDL (RR= 1.01, 95%CI (0.84, 1.22)) and high triglycerides (RR= 0.95, 95%CI (0.81, 1.10)) on head and neck combined cancer (**Table 2**).

An apparent obesity paradox demonstrated in individual studies ^55,67^ might be explained by BMI as a collider acting as a form of selection bias ^68^. While tobacco smoking is correlated with obesity, this may be dose dependant and change following smoking cessation ^69^.

Furthermore, many HNC patients are more likely to be underweight at presentation, particularly those with alcohol use disorders, often correlated with heavy smoking behaviour. Variability in the results may exist due to differences in timing of the exposure measurement, whereby misclassification of participants into the wrong BMI category could have occurred. This is because around one-third to a half of HNC patients experience between 5 – 10% weight loss during the period prior to diagnosis ^70^. Obesity may also provide additional nutritional reserves giving patients a survival advantage, if for example there was latent tumour progression prior to the baseline diagnosis in these observational studies. There may be issues with measurement error again leading to misclassification in this study, given BMI is simply a function of mass and height and does not specifically indicate metabolically unhealthy adiposity or take into account lean muscle mass. In future, more accurate measures body composition such as bioelectrical impedance or dual-energy X-ray absorptiometry (DEXA) scanning could be used to better differentiate fat mass from lean tissue. This study also lacked detail on HPV status or tumour stage, with more advanced stage disease likely resulting in more significant weight loss ^71^. Subgroup differences were found across metabolic traits with respect to geographic location. Stronger effects were found for obesity in USA compared with the UK, and for both type 2 diabetes and dyslipidaemia in Asian populations compared with the other continents, which is in keeping with global epidemiology ^72,73^. Larger subgroup effects were also found when patient’s self-reported their diagnoses compared to researchers using medical records, suggesting recall bias may be present.

### Strengths and limitations of this study

This meta-analysis includes the largest available studies, resulting in a sample size of n= 39,002 HNC cases in a total population size of over 84 million. This suggests the study was well powered to detect any association between metabolic traits and HNC, however there were fewer studies for hypertension and dyslipidaemia compared to the other traits. As there were <10 studies in these analysis, a comprehensive assessment of publication or reporting bias could not be made. However, the lack of smaller positive or negative effect studies for type 2 diabetes suggests potential publication or reporting bias. Both the obesity and type 2 diabetes funnel plots revealed outlier results. Leaving out the two outlier studies in the obesity meta-analysis had little effect on the overall null finding. Evidence of substantial heterogeneity was present (I^2^ values >50%) between studies across all metabolic traits, which increased when combining HNC subsites as would be expected. In particular for type 2 diabetes whereby four out of five outlier studies were in Asian only populations. This could be due to the differences in HNC aetiology in Asia e.g., betel quid chewing. Future work should include meta-analysing studies separately by geographic region or restricting to particular populations, as this appears to be a clear source of heterogeneity. Other forms of bias such as collider, recall and information bias may have been present, as described in the previous section. Despite multivariable adjustment within studies, residual confounding may still exist, given that HNC risk factors, such as smoking, alcohol and socioeconomic status are difficult to fully account for. While we conducted a number of subgroup analyses, there was insufficient data in the included studies to examine the effect of sex. This may have been important as previous analyses have suggested that the effect of type 2 diabetes on HNC was mainly found in females ^17^, which may indicate poorer metabolic control, elevated exposure to insulin and subsequent oxidative DNA damage. There were limited data for smoking at specific subsites across the metabolic traits and therefore meta-analysis at this level was not possible.

## Conclusions

The meta-analysis performed in this study suggested there was limited evidence for an association between obesity or type 2 diabetes and HNC risk. However, there was some evidence for increased risk associations with hypertension and dyslipidaemia. These observational associations are susceptible to confounding, bias and reverse causality and results must be interpreted with caution given the high risk of bias detected.

## Authors’ contributions

AG and CR jointly contributed to the development of the protocol, and the drafting, writing and editing of this manuscript. FS contributed to the development of this study and editing of the manuscript. JH contributed to the development of the search strategy. BM, EG, EV, RR and JPTH contributed to the development of this study. MG was responsible for conceptualising this study, drafting and editing this manuscript. All authors contributed to the development of the search strategy. All authors have approved and contributed to the final written manuscript.

## Patient and Public Involvement

Patients and/or the public were not involved in the development, conduct or proposed dissemination of this systematic review protocol.

## Data Availability

All data produced in the present work are contained in the manuscript.

## Supplementary information

Supplementary Appendix 1 Systematic review protocol

Supplementary Appendix 2 PRISMA checklist

---

**Supplementary Table 1.**
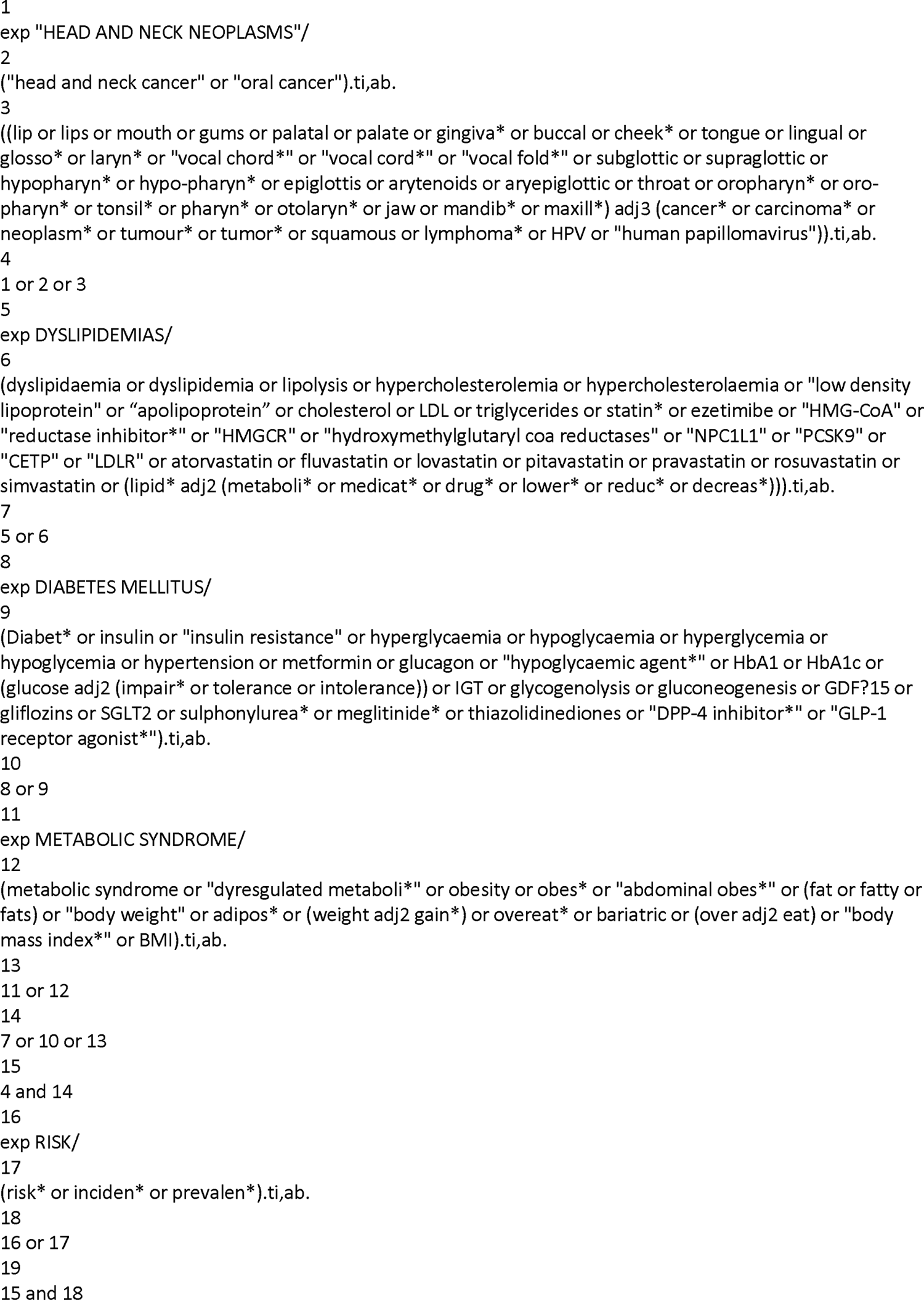
Search strategy.

**Supplementary Table 2.**
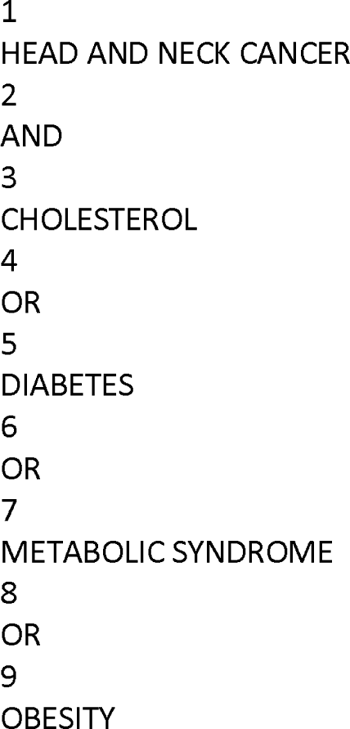
Modified search strategy.

**Supplementary Table 3.**
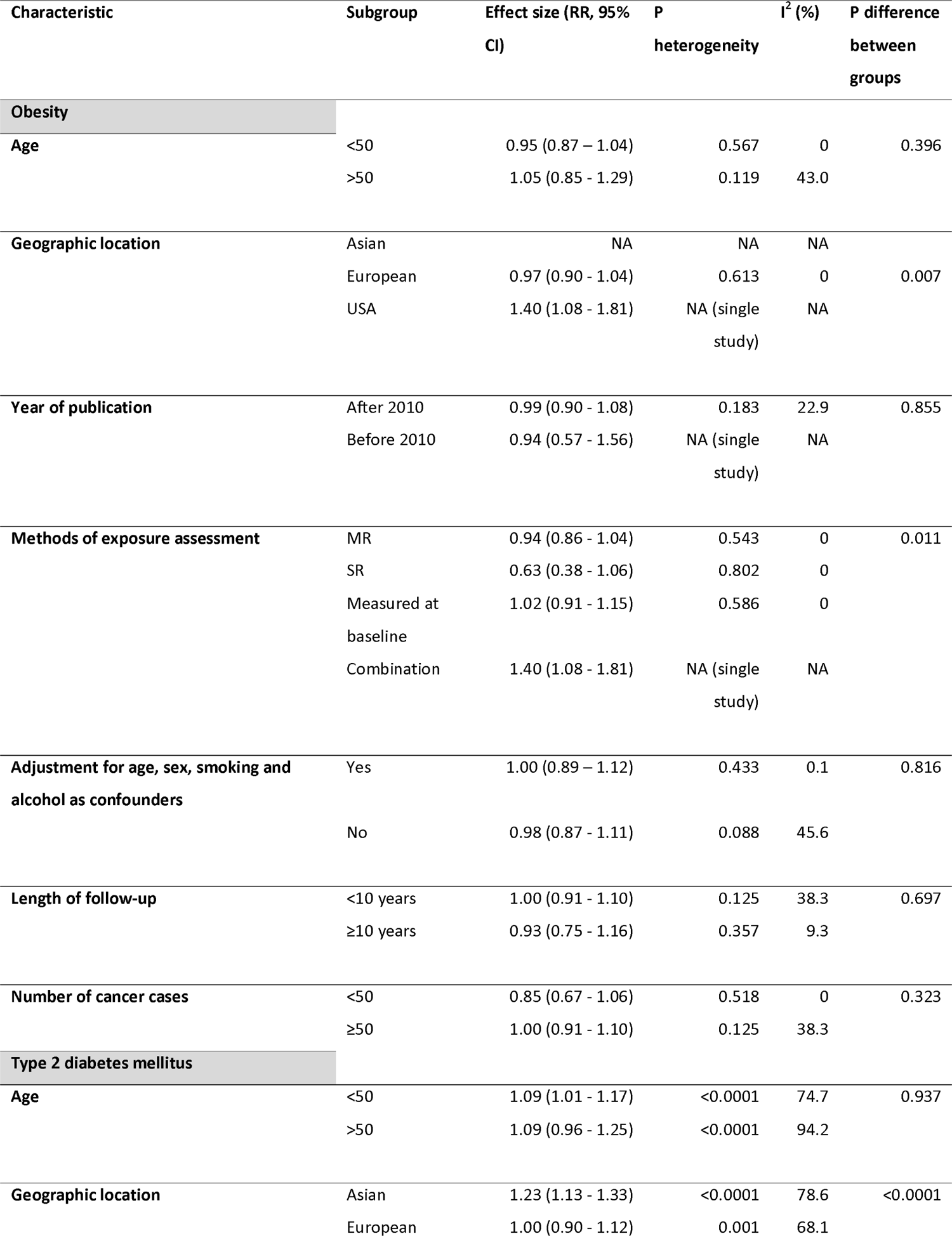
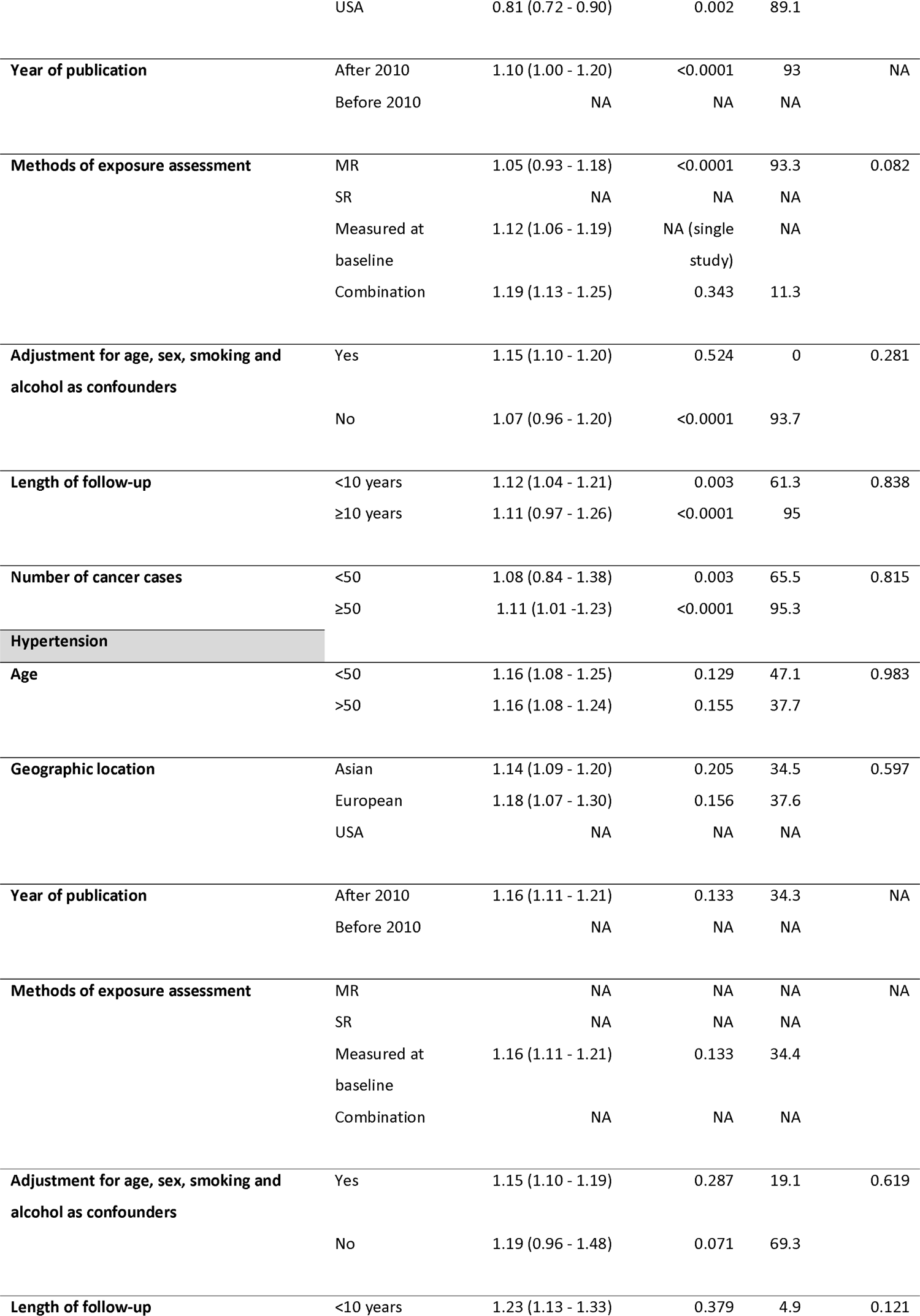
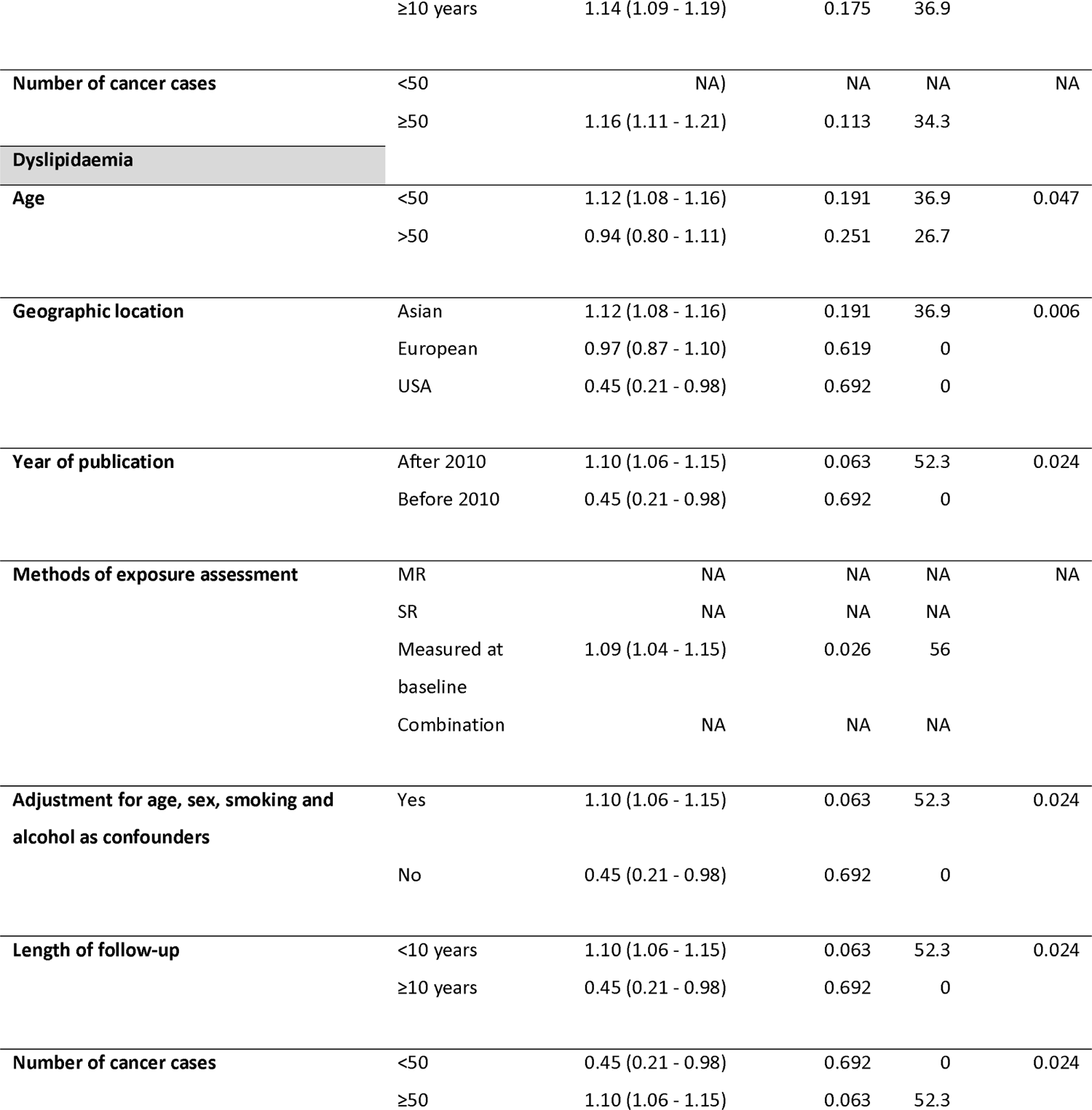
Subgroup analyses for the association of metabolic disorders and risk of head and neck cancer.

Supplementary Figures 1 – 4 Risk of bias assessments for each metabolic trait using the ROBINS-E tool

**Supplementary Figure 1.**
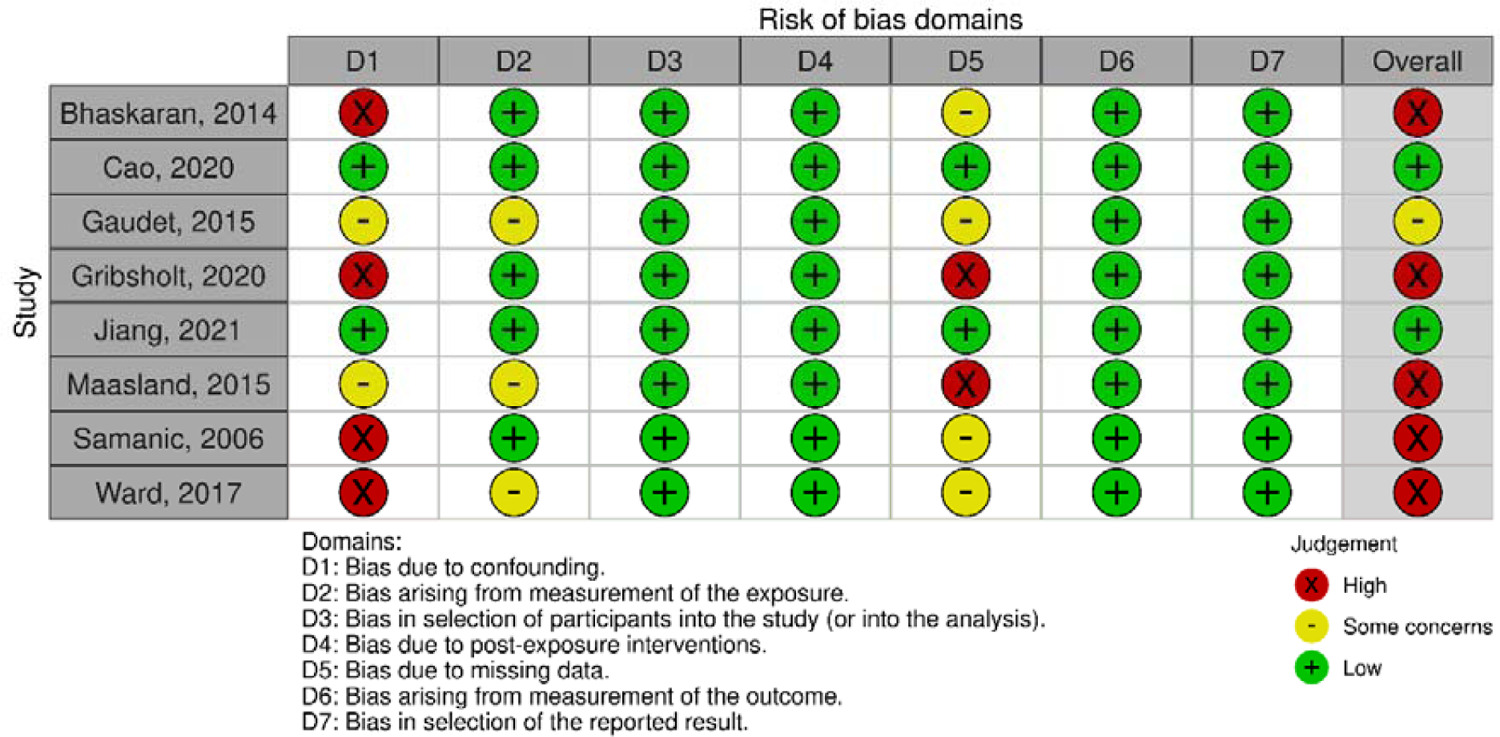
Obesity

**Supplementary Figure 2.**
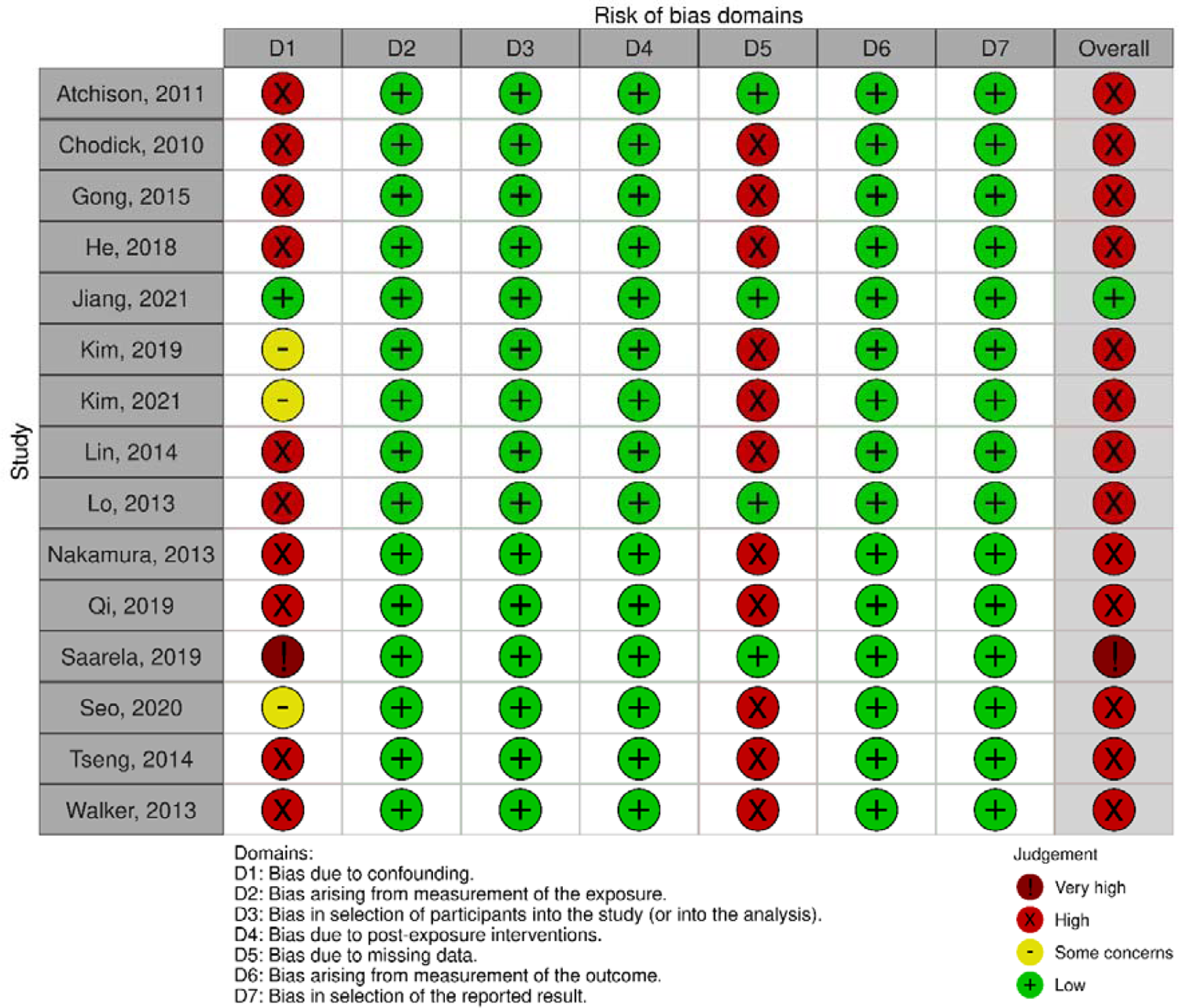
Type 2 Diabetes

**Supplementary Figure 3.**
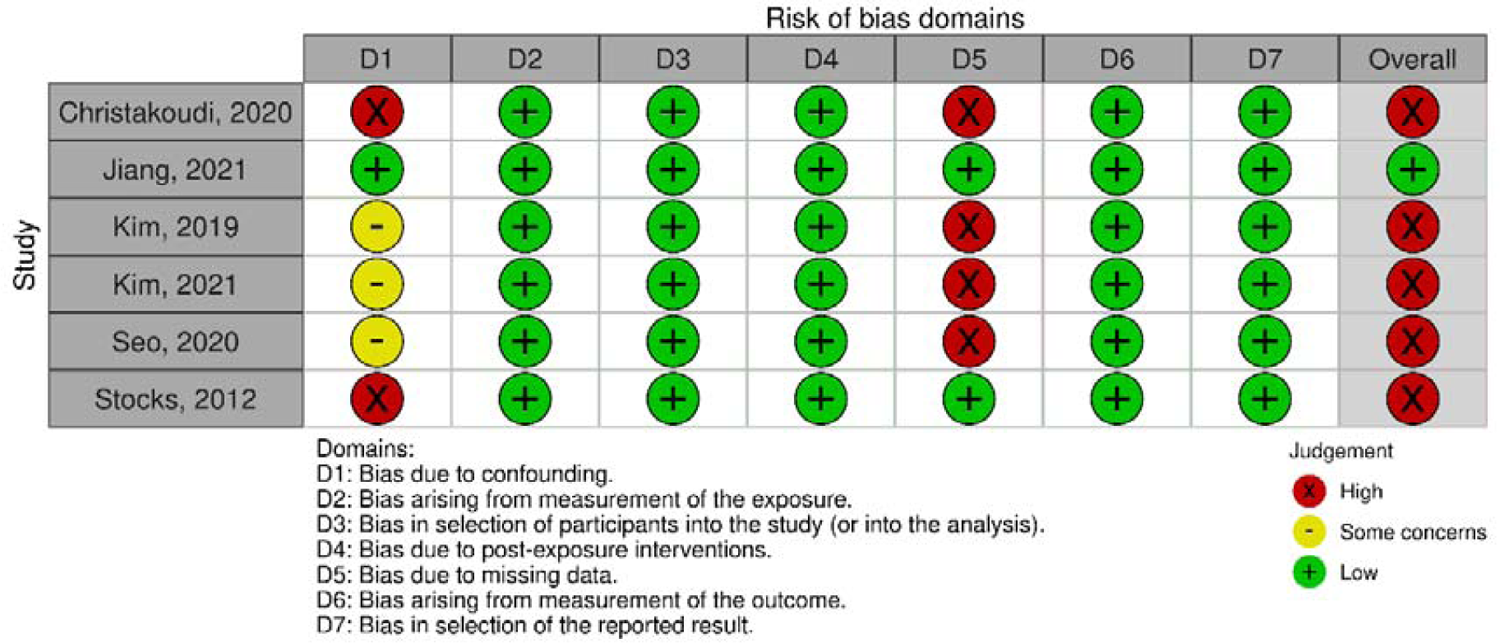
Hypertension

**Supplementary Figure 4.**
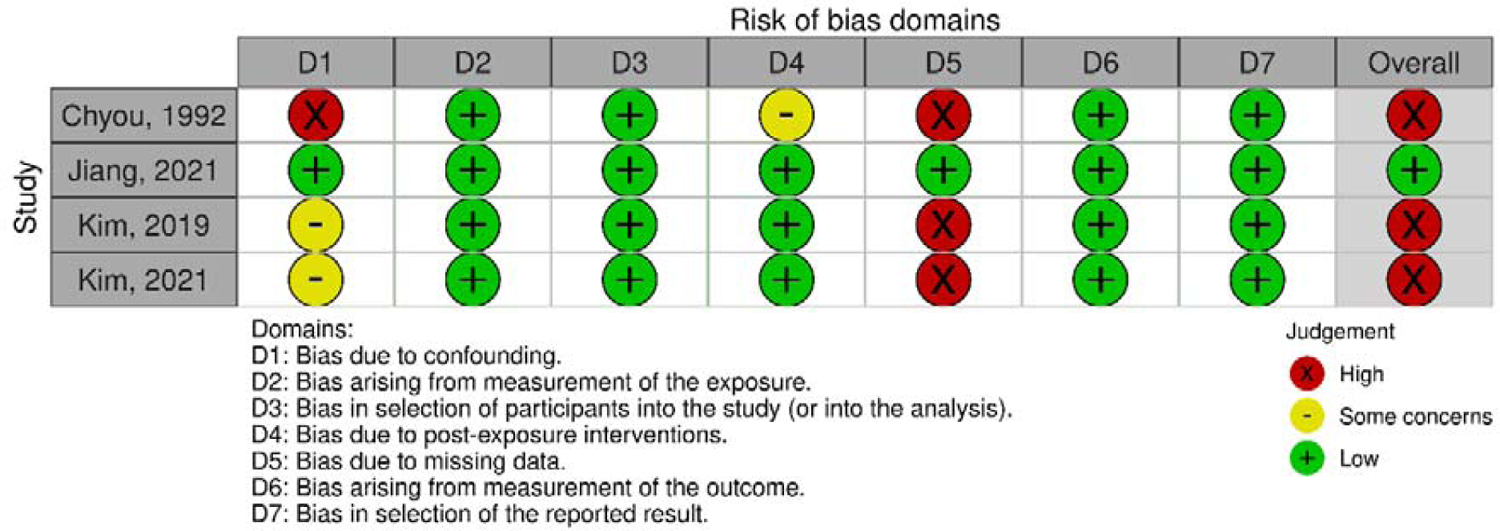
Dyslipidaemia

